# Predicting the effects of COVID-19 related interventions in urban settings by combining activity-based modelling, agent-based simulation, and mobile phone data

**DOI:** 10.1101/2021.02.27.21252583

**Authors:** Sebastian A. Müller, Michael Balmer, William Charlton, Ricardo Ewert, Andreas Neumann, Christian Rakow, Tilmann Schlenther, Kai Nagel

## Abstract

Epidemiological simulations as a method are used to better understand and predict the spreading of infectious diseases, for example of COVID-19.

This paper presents an approach that combines a well-established approach from transportation modelling that uses person-centric data-driven human mobility modelling with a mechanistic infection model and a person-centric disease progression model. The model includes the consequences of different room sizes, air exchange rates, disease import, changed activity participation rates over time (coming from mobility data), masks, indoors vs. outdoors leisure activities, and of contact tracing. The model is validated against the infection dynamics in Berlin (Germany).

The model can be used to understand the contributions of different activity types to the infection dynamics over time. The model predicts the effects of contact reductions, school closures/vacations, masks, or the effect of moving leisure activities from outdoors to indoors in fall, and is thus able to quantitatively predict the consequences of interventions. It is shown that these effects are best given as additive changes of the reinfection rate R. The model also explains why contact reductions have decreasing marginal returns, i.e. the first 50% of contact reductions have considerably more effect than the second 50%.

Our work shows that is is possible to build detailed epidemiological simulations from microscopic mobility models relatively quickly. They can be used to investigate mechanical aspects of the dynamics, such as the transmission from political decisions via human behavior to infections, consequences of different lockdown measures, or consequences of wearing masks in certain situations. The results can be used to inform political decisions.

**Author summary:** Evidently, there is an interest in models that are able to predict the effect of interventions in the face of pandemic diseases. The so-called compartmental models have difficulties to include effects that stem from spatial, demographic or temporal inhomongeneities. Person-centric models, often using social contact matrices, are difficult and time-consuming to build up. In the present paper, we describe how we built a largely data-driven person-centric infection model within less than a month when COVID-19 took hold in Germany. The model is based on our extensive experience with mobility modelling, and a synthetic data pipeline that starts with mobile phone data, while taking the infection dynamics and the disease progression from the literature. The approach makes the model portable to all places that have similar so-called activity-based models of travel in place, which are many places world-wide, and the number is continuously increasing. The model has been used since its inception to regularly advise the German government on expected consequences of interventions.

## Introduction

When COVID-19 took hold in Germany in February 2020, there was an urgent need for a differentiated modelling capability to predict the consequences of interventions. We used our decades-long experience with person-centric modelling of traffic [1] to build a first prototype within two weeks [2]. An advantage of using this starting point is that the whereabouts of all simulated persons, including their overlapping time spent at facilities or in (public transport) vehicles, are already given by the model, which is derived in part from mobile phone data. Since the input data contains age as an attribute of each synthetic person, it was straightforward to include agent-dependent disease progression into the model from the start.

The first version of our model used uniform contact intensities everywhere, and in consequence predicted infections mostly where people spend time with other people. As a result, that model predicted that a 50% reduction in leisure activities would reduce the reinfection rate R by 15% while a 50% reduction in work activities would reduce R by 19%. A reduction of all out-of-home activities by 50% resulted in a reduction of R by 62% [3].

We also could use our data source to provide the average daily duration of out-of-home activities since March. Once they went down, we used that as a data feed for our model to reduce the number of out-of-home activities accordingly. The model was then improved to include the effect of aerosol transmission [4], by taking facility size and air exchange rates into account, different for each activity type [5]. We also included lower susceptibility and infectiousness of children as was by then reported in the literature [6, 7]. Based on that model, we predicted that the re-opening of the schools in Berlin after the summer vacation would be noticeable in the infection numbers, but it would not push the infection dynamics into criticality [8]. We also predicted a second wave in fall, based on a model for performing leisure activities indoors vs outdoors, with infection probabilities reduced by a factor of 10 in outdoors conditions [4, 9]. After that second wave came earlier than predicted, we re-evaluated our assumptions about spring vs. fall temperature sensitivity; having people move outdoors at temperatures above 17.5C in spring while having them move indoors at temperatures below 25C in fall now explains the dynamics of the epidemics in Berlin quite well.

The model is regularly used to advise the German federal government (e.g. [10, 11]). The current main contribution of those reports is to provide differentiated predictions of the influence of various interventions, such as reductions of activity participation, masks, or vaccinations. For the present paper, we show the contributions of different activity types to the infection dynamics as predicted by the model. We show how most activity types generate over time fairly constant contributions to the reinfection rate R, and in consequence it is structurally more stable to report reductions of R caused by interventions as an additive than a multiplicative term as is usually done (e.g. [12]). The model also explains why there are decreasing marginal returns to stay-at-home interventions [13]. Finally, the model makes a prediction concerning the magnitude of the difference between summer and winter, caused by moving activities indoors during winter.

## Related work

### Location information from mobile device data

There are two general methods to obtain location information from mobile phone data (cf. [14]):

- **Network-based** means that the network operator has some knowledge about the location of a switched-on mobile device. There are several techniques, which depend on the technical equipment of the network infrastructure, and in part also on corresponding infrastructure on the mobile devices. In general, the operator keeps the so-called call detail records (CDRs), which state, for each data or voice call, the time and the cell tower ID. Often, operators also keep cell tower handovers even when no calls are made. More spatial precision can be obtained by sending corresponding requests to the mobile device, but this is not done by default and thus in general not available. The network-based approach can keep trajectories; in principle from switching on to switching off of the mobile device; in practice often limited by privacy laws, in Germany for example to 8 hours after which the temporary ID of a cell phone needs to be changed without connecting it to the previous ID.
- **Device-based** means that the mobile device itself determines its position and sends it to the data collector. A typical example are map applications, which collect and send GPS positions while the application is running. Other examples are weather apps, or in general all apps which contain location-providing libraries in their code. Many mobile devices inform the user about the fact that location information is given away, but some do not. A special case are apps that are specifically about collecting movement data, such as [15]. The device-based approach is spatially more precise. It is, on the other hand, limited to those devices that use the particular app or library. In consequence, only a small subset of devices sends this data, and only from starting to ending the particular app.

As a result, the device-based approach is very good for providing crowdedness data, which is, for example, provided by google for many facilities, and which is also the basis for google mobility reports [16]. In order to obtain longer trajectories for large population samples, the network-based approach is better.

Mobile device data can be used in many ways to help with analyzing the dynamics of an epidemics. Out of the categories described by Grantz et al. [14], the work described in this paper concentrates on “Capturing epidemiologically-relevant behaviors with mobile phone data”. Note that there are two separate, but related inputs where we use mobile device data:

1. To obtain the “regular” movement patterns of our synthetic population.
2. To obtain the reductions of those movement patterns throughout the unfolding of COVID-19 in Germany.

### Daily activity trajectories

Using daily activity chains as the basis for transport modelling is an established approach in the transport modelling community (e.g. [17–19]). An activity chain is a sequence of activities of a person, where activities have types such as home, work, shop, etc., starting and ending times, and locations. There are several ways to generate such activity chains, for example by using activity-based demand generation models (e.g. [18, 20–23]), by taking them from travel diaries (e.g. [24, 25]), by using mobile phone data (e.g. [26]), or by data fusion from open access data sources (e.g. [27]).

In the present situation, we needed a technology that was readily available, allowed uniform rollout at least in Germany, and that would allow to be updated by changes in the mobility behavior during the unfolding of the COVID epidemics. For that reason, we used an established process that generates activity chains mostly from mobile phone data [26]. The process is described in more detail in S1 Text. The outcome of the process are activity chains, encoded as events (cf. Fig. 1), for as many synthetic persons as Germany has inhabitants. Since the activity chains stem from transport modelling, they also contain knowledge about trips between activities, importantly trips by public transport, and in consequence also contain, for each synthetic person, events when they enter or leave certain public transit vehicles.

**Fig 1.**
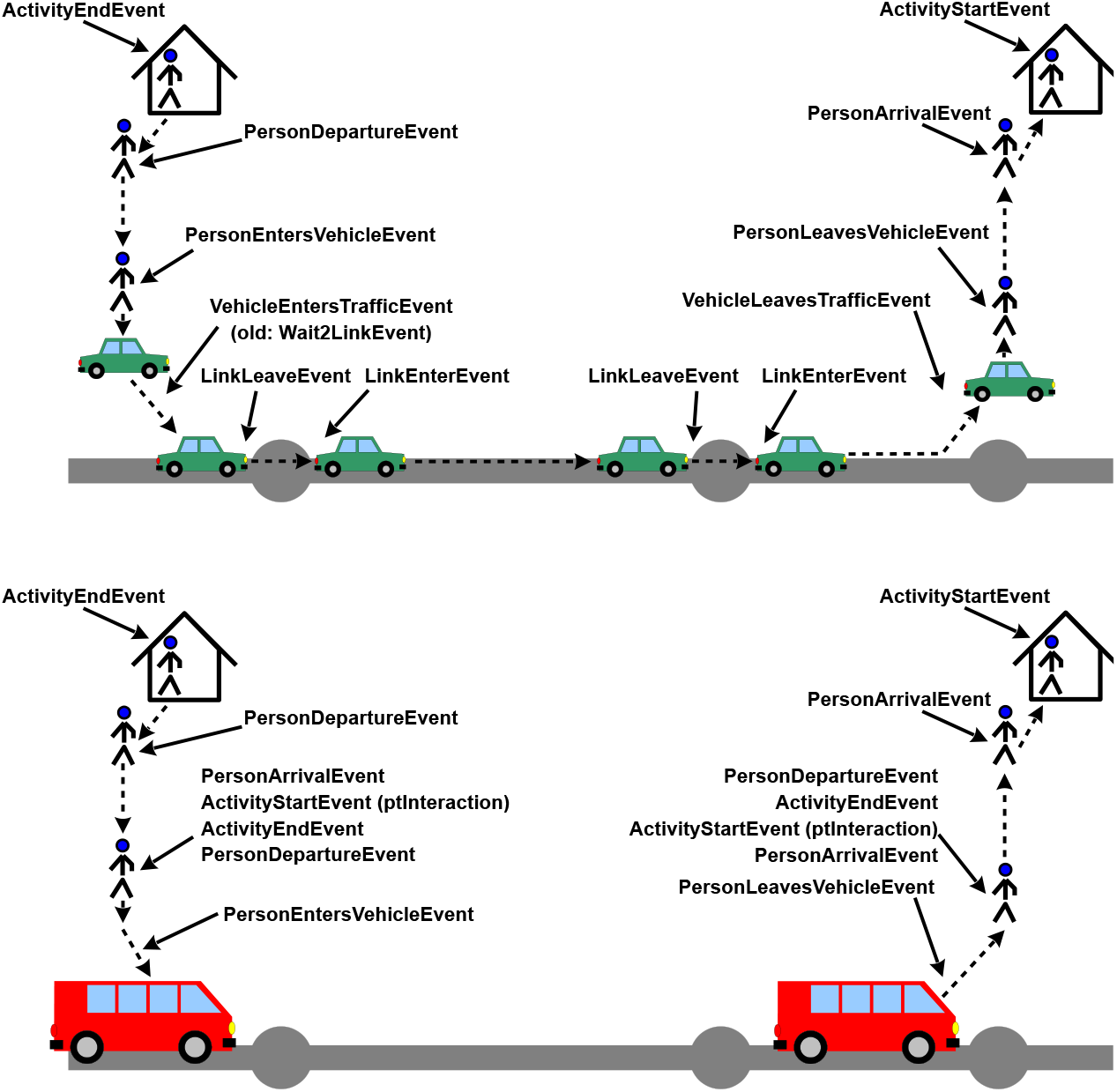
TOP: Events for travel by individual vehicle. BOTTOM: Events for travel by public transport. Source: [1].

### Using mobile device data to observe changes of mobility behavior during COVID-19

There are many studies that look at how mobility has changed during the unfolding of the COVID pandemic.

Apple [28] and Google [16] both publish mobility data based on usage data from their map services. The Apple data shows for different countries and cities how the use of different transport modes has changed over time. Different from Apple, Google publishes mobility data with a focus on activity participation per activity type. This makes it possible to evaluate how the population has reacted to restrictions (e.g. to what extent leisure or work activities were reduced).

Warren et al. [29] use mobile device location data to measure distances travelled by the population in the investigated period from January 2020 until March 2020. Their analyses show that for several U.S. states and counties, significant reductions in mobility can be detected, especially from mid-March onward.

Pullano et al. [30] use travel flow data derived from mobile phone data from France to evaluate the impact of a lockdown which was implemented in France in mid March. One result of their study is that on a nationwide level, trips were reduced by about 65%.

Bonaccorsi et al. [31] use Italian mobility data to analyse how mobility is linked to economical parameters. They find that mobility was reduced stronger during the lockdown for cities with a low income per person and that financial help from the government is needed to prevent an increase in poverty.

Eisenmann et al. [32] use a representative travel survey in Germany to find out whether the pandemic has influenced transport mode choice. Their results show that use of individual modes like car have increased during the lockdown in April whereas the use of public transport has declined.

Axhausen et al. [33] conduct a study to record mobility behavior using a smartphone app to track the impact of various measures during the pandemic. Their regularly published reports include, for example, information on how miles traveled by mode of transportation change over time, how trip purpose changes, or how trip durations change.

### From reductions of mobility behavior to reductions of infections

Describing mobility changes “during Corona” is, however, not our primary focus; rather, we are interested in how the infection dynamics can be better understood and possibly predicted with the help of mobility and other data.

A possible approach to achieve this is data mining; for example, Badr et al. [34] correlate a mobility ratio *MR* with a COVID-19 growth ratio *GR*. They find a strong correlation, as may be expected. However, the correlation coefficient between *MR* and *GR* changes over time (see Fig. 3 of their paper), implying that the dynamics is more complex than what can be captured by that correlation.

Fritz and Kauermann [35] use anonymized Facebook movement data to investigate the relation between COVID infections and movements; they find, together with other effects, that both reduced mobility and reduced diversity in visits reduces infection numbers. Again, they allow separate coefficients for each week, which is different from what we are aiming for.

Jia et al. [36] and Xiong et al. [37] look at how long distance travel influences the disease import; they find that a high inflow from areas with high incidences is positively correlated with high infection numbers. They do not, however, look at disease spread within the urban fabric, driven by daily movement patterns.

Lau et al. [38] fit a spatio-temporal transmission process model, based on [39], to individual infection surveillance data. The model uses, between other elements, an exponential distribution for an infection to jump to a new location; the mean distance of that distribution is taken from Facebook mobility data, reduced accordingly after the stay-at-home order is introduced, and in consequence reducing infections. The model does not include different infection contexts, and is in general in its present form more suitable for rural contexts, where spatial effects play a larger role than in urban contexts.

Fairly close to our work are Chang et al. [40]. They first construct, based on mobile phone data, a mobility network between census block groups and points of interest based on mobile phone data, and then use that model to investigate reopening strategies. The paper does not state it explicitly, but the data presumably comes from the device-based approach. In consequence, they have a very detailed resolution of the facilities (they differentiate, e.g., between full-service restaurants, limited-service restaurants, and cafes/snack bars), but on the other hand they do not simulate individual synthetic persons. We will come back to similarities and differences in our discussion.

### Person-centric epidemiological modelling

The general dynamics of virus spreading is captured by compartmental models, most famously the so-called SIR model, with *S* = *susceptible, I* = *infected/infectious*, and *R* = *recovered* [41, 42]. Every time a *susceptible* and an *infectious* person meet, there is a probability that the susceptible person becomes infected. Some time after the infection, the person typically recovers. Variants include, e.g., an *exposed* (but not yet *infectious*) compartment between *S* and *I*.

Instead of running these models with compartments, one can run them on a graph [43–47]. Persons are represented as vertices, connections between persons are denoted as edges. The random interactions that are implied by the compartmental models are then replaced by interactions with graph neighbors.

In reality, these interactions change from day to day; in particular, possible superspreading events like weddings or other large gatherings cannot be encoded in a static graph. For this, temporal networks have been investigated ([47], section VIII).

Finally, a “different framework emerges if we consider nodes as entities where multiple individuals or particles can be located and eventually wander by moving along the links connecting the nodes” [47]. When COVID-19 took hold in Europe, a model of this type by Imperial College [48, 49] had a large impact on policy in the UK. Other examples of this approach are by the Virginia Biotechnology Institute [48, 50, 51] and by the Center for Statistics and Quantitative Infectious Diseases in Seattle [48, 52]. Examples for similar approaches on the global or regional level are [43, 53–55]. Groups that started more recently include [56] and [57].

Aleta et al. [58] construct an agent-based model, similar to ours. Their data derives from the device-based approach, as explained above, from persons specifically recruited to collect their long-term trajectories. In consequence, they have long trajectories, with high spatial precision, but for only 2% of the population (which is still an impressive sample). We come back to this in our discussion.

A special case is by Kucharski et al. [59], who use a pre-existing dataset with recorded social contacts for 40 162 participants. This is close to our approach in that the persons who encounter each other for how long and in which context are microscopically specified. Differences include that it is not a model for the full population of a region, and the study does not trace behavioral changes throughout the pandemic.

### Person-centric epidemiological models derived from transport simulations

Smieszek et al. [60, 61] and Hackl and Dubernet [62] constructed epidemiological models on top of pre-existing transport simulations; these are the main starting point for us.

Najmi et al. [63] start from a person-centric transportation planning model for Sydney, and add a disease transmission model that computes possible infections based on co-locations during the simulated day. The approach is similar to ours, but does not use mobile phone data to track the actual mobility behavior. They also do not use an infection model that depends on the spatial situation of the activity type.

The works by Manout and Ciari [64, 65], for Montreal, and by Bossert et al. [66], for South Africa, are based on an earlier version of the model used in the present paper.

Except for Chang et al. [40], we have not found other studies that use mobile phone data both to generate the base mobility *and* to adjust the mobility throughout the unfolding of the epidemics.

## Model details

Important sub-models of agent-based epidemics models are: contact model, infection model, and disease progression model. These are described in more detail in the following sections.

### Mobility model and resulting contact model

As stated, we take the synthetic persons and their movements from transport modelling, cf. Fig. 1. For the present study, the data is generated by a synthetic method developed by Senozon, detailed in Sec. S1 Text. We have used and are using the same data for other projects [67–70]. From these activity chains, we extract how much time people spend with other people at activities or in (public transport) vehicles. That is, infection opportunities are directly taken from the input data. Details are provided in Sec. S2 Text.

Importantly, we also take the *reductions* of activity participation over time from mobility data. The method is the same as for the generation of the activity patterns but stopped early, cf. Sec. S1 Text. Fig. 2 shows, in blue, the percent reduction of all out-of-home activities since 2020-03-01; weekly averages of these data are used as input to the model. Sec. Reductions of activity participation in the appendix provides details.

**Fig 2.**
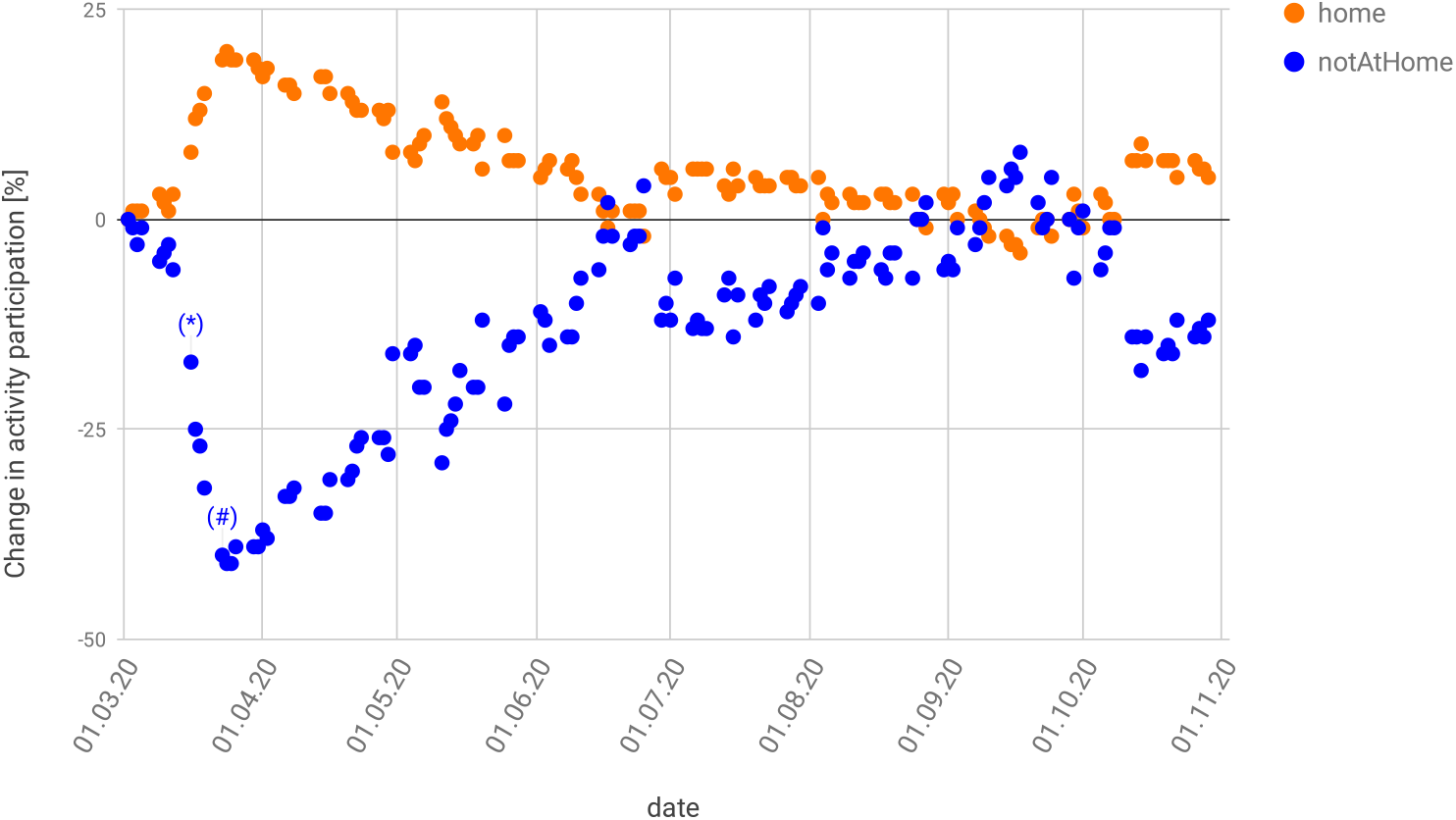
Change in activity participation compared to the baseline for normal workdays. All out-of-home activities are combined into one number. (*) denotes the first day of closures of schools, clubs, and bars; and (#) the first day of the so-called contact ban which came together with closures of all restaurants and non-essential stores.

### Infection model

Once two persons are identified to have contact, and one of them is contagious and the other is susceptible, there is a probability of an infection. For this, we use the mechanical model by Smieszek [60, 71]: Infected persons generate a “viral load” that they exhale, cough or sneeze into the environment, and people close by are exposed. Overall, the probability for person *n* to become infected by this process in a time step *t* is described as

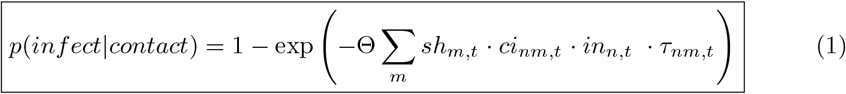

where *m* is a sum over all other persons, *sh* is the shedding rate (∼ microbial load), *ci* the contact intensity, *in* the intake (reduced, e.g., by a mask), *τ* the duration of interaction between the two individuals, and Θ a calibration parameter.

For small values of the exponent, one can approximate Eq. (1) as

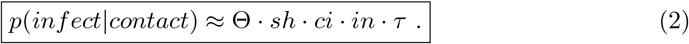

We do not use this approximation in our computer implementation, but it helps understanding the following arguments.

All parameters can be given in arbitrary units as long as they are always the same since the units are absorbed by Θ.

### Contact intensities

For SARS-CoV-2, it is plausible to assume that a large share of the virus material is shed as aerosol [4]. In consequence, the first relevant term to compute the viral concentration in the air is the shedding rate, *sh*.

For such aerosols, it is plausible to assume that they mix quickly into the room, leading to the same uniform concentration everywhere [72]. Evidently, that concentration is indirectly proportional to room size: if the room is twice as large, the resulting concentration is half as large.

Next, air exchange plays a role [72]. One could, for example, assume that the windows are opened once per hour, and all of the air is replaced with outside air. This would correspond to an air exchange rate of 1/h. If one assumes a constant rate of virus emission, there would be a linear increase of concentration up to the opening of the window, after which (in a theoretical model) the virus concentration in the air would quickly drop to zero. The *average* virus concentration over this process would be half as much as the maximum concentration just before window opening. In consequence, the resulting average concentration is indirectly proportional to the air exchange rate: If the air is exchanged twice as often, the resulting average virus concentration is half as large. This also holds for continuous air exchange, e.g. by mechanical means.

All of the above together replaces Eq. 2 by

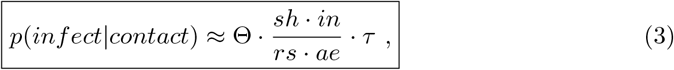

where *rs* is the size of the room, and *ae* is the air exchange rate. That is, it sets

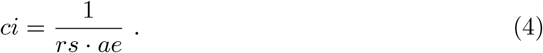

Again, the physical units are absorbed into Θ; note, however, that the air exchange rate *ae* is defined as exchanging air for the full room, and not in, say, cubic meters.

### Estimation of room sizes

As stated above, our data resolves down to the level of “facilities”. These correspond roughly to buildings. In consequence, such a facility can be anything from a single family home to a large office building to a sports arena.

Since our simulation tracks when persons are at facilities, we can, for each facility, obtain the maximum number of persons at that facility, 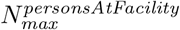, over the day.

In addition, one can obtain typical floor space per person, *fs*, from regulatory norms and other sources (see Tab. 1). This leads to

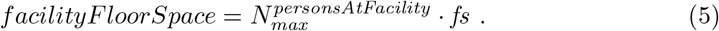

**Table 1.** Air volume flows per person used in the model. The floor area per person and the air exchange rate come from building manuals or similar standards. The share of old buildings/vehicles is an estimate. Universities are assumed to have twice as much space per student as schools. Shop, errands, and business are assumed to follow the same characteristics as work. The average of the volume flow per person is computed as 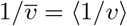.^*a*^

| activity type | area per person<br>[m<sup>2</sup>] | air ex-<br>change<br>rate low<br>[1/h] | air ex-<br>change<br>rate high<br>[1/h] | volume<br>flow per<br>pers. low<br>[m<sup>3</sup>/hr/p] | volume<br>flow per<br>pers. high<br>[m<sup>3</sup>/hr/p] | share old<br>buildings<br>/vehicles | resulting<br>volume flow<br>per pers.<br>[m<sup>3</sup>/hr/p] |
| --- | --- | --- | --- | --- | --- | --- | --- |
| home [74] | 22 | 0.5 | 0.5 | 33 | 33 |  | 33 |
| schools, day care [75] | 2 | 0.5 | 0.5 | 6 | 6 | 100% | 3 |
| universities | 4 | 0.5 | 0.5 | 12 | 12 | 100% | 6 |
| public transport | 0.33 | 2.0 | 10.0 | 1.98 | 9.90 | 50% | 3.3 |
| leisure [76] | 1.25 | 0.5 | 10.0 | 1.88 | 37.50 | 50% | 3.57 |
| shop | 10 | 0.5 | 1.5 | 15 | 45 | 10% | 37.5 |
| work [77–79] | 10 | 0.5 | 1.5 | 15 | 45 | 50% | 22.5 |
| errands | 10 | 0.5 | 1.5 | 15 | 45 | 50% | 22.5 |
| business | 10 | 0.5 | 1.5 | 15 | 45 | 50% | 22.5 |

<sup>a</sup>Averaging the *inverse* of the volume flows is the correct approach since the contact intensities need to be averaged. Assume, say that with probability 1/2 one enters either a facility with $v = 1$ or with $v = \infty$ . Then the resulting probability to become infected is half of that if entering the room with $v = 1$ with probability one. This corresponds to $v = 2$ , meaning that the inverse of the $v$ need to be averaged.

Since we divide all facilities by *N*^*spacesPerFacility*^ (cf. Sec. Handling of large facilities in the appendix), this leads for the room size to

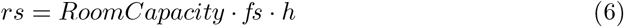

where *h* is the room height, and

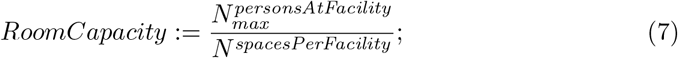

note that *N*^*spacesPerFacility*^ = 1 for home activities (cf. Sec. Handling of large facilities in the appendix).

### Air exchange

Overall, the above results in

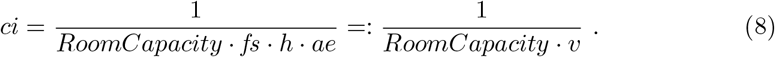

*v* = *fs · h · ae* is the air volume flow per person at capacity, which is also used by other models as the decisive quantity to decide about the relatively safety of rooms [5].

*RoomCapacity* describes the size of the room; a room for a certain function (e.g. “work”) typically comes with a certain *v* which is per person; this is then multiplied by *RoomCapacity* which gives the air exchange volume flow for the room. Since these equations calculate the conditional infection probability *given that there is one infectious person in the room*, it is clear that a larger room under these conditions results in a smaller infection probability.

However, if the room is twice as large, then there will presumably also be twice as many persons in it, doubling our own risk, and thus in the average cancelling out the effect of the larger room size. This second effect, however, is computed directly by our contact model (Sec. Mobility model and resulting contact model above), and thus does not have to be included into the conditional infection probability. This has the additional advantage that if a person is in large container outside its peak usage, the model will calculate a much reduced infection probability. Examples for this are public transport vehicles, premises for large events, or restaurants.

Table 1 gives concrete values that the model presented in this paper uses. Kriegel [73] currently recommends an air volume flow per person of at least 75 *m*^3^*/h* to suppress most COVID infections. This corresponds well with the values of Table 1, which state that average facilities, when operated at full occupancy, are in general not “safe”.

### Children

Current research implies that the susceptibility and infectivity are reduced for children compared to adults. We model this by including the susceptibility and infectivity into Eq. (1). For adults both parameters are set to one. For people below the age of twenty the infectivity is reduced to 0.85 and the susceptibility to 0.45 [6, 7]. Note that this does not mean that the infection probability for children is necessarily lower than for adults because children are more likely to perform activities with a high contact intensity (= low air volume flow per person), as shown in Table 1.

### Disease progression model

The disease progression model is taken from the literature [80–85] (also see [86]). The model has states *exposed, infectious, showing symptoms, seriously sick* (= should be in hospital), *critical* (= needs intensive care), and *recovered*. The durations from one state to the next follow log-normal distributions; see Fig. 3 (LEFT) for details. We use similar age-dependent transition probabilities as [49], shown in Fig. 3 (RIGHT).

**Fig 3.**
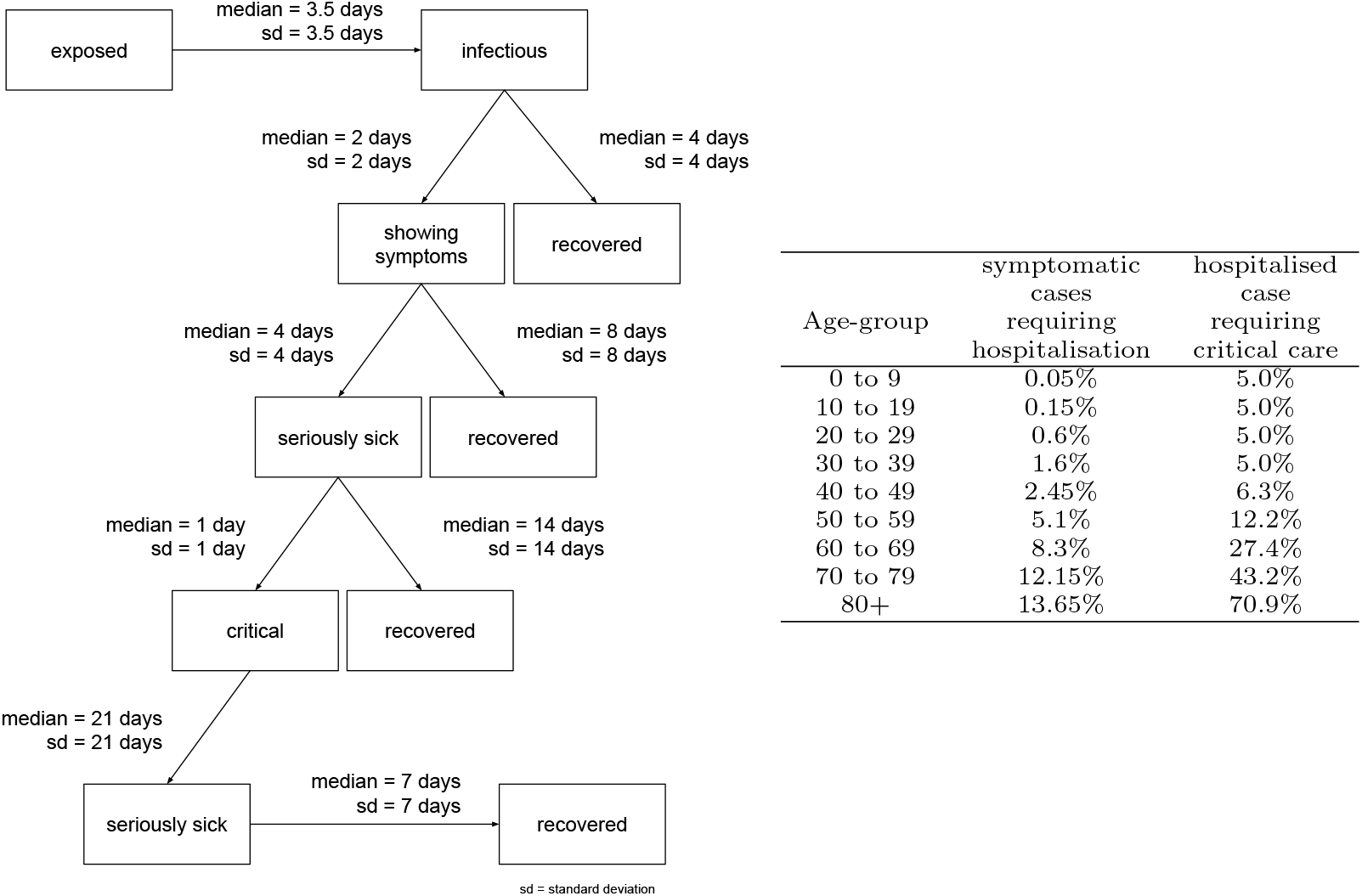
LEFT: State transitions [80–85]. RIGHT: Age-dependent transition probabilities from symptomatic to seriously sick (= requiring hospitalisation), and from seriously sick to critical (= requiring breathing support or intensive care). Source: [49], except that the numbers in the second column are divided by 2 (discussed in Sec. Under-reporting).

Infecting another person is possible during *infectious*, and while *showing symptoms*, but no longer than 4 days after becoming *infectious*. This models that persons are mostly infectious relatively early through the disease [81], while in later stages the infection may move to the lung [82], which makes it worse for the infected person, but seems to make it less infectious to other persons.

### Simulation runs

This paper presents simulation results for the metropolitan area of Berlin in Germany, with approx. 5 million people. A typical simulation run looks as follows:

1. One or more *exposed* persons are introduced into the population.
2. At some point, *exposed* persons become *infectious*. From then on, every time they spend time together with some other person in a vehicle or at some activity, Eq. (1) is used to calculate the probability that the other person, if *susceptible*, can become infected (= *exposed*). If infection happens, the newly infected person will follow the same progression.
3. *Infectious* persons eventually move on to other states, as described in Fig. 3.

The model runs many days, until no more infections occur.

## Methods and results

### Calibration

The calibration procedure undertaken for the present paper is described in Sec. Appendix: Calibration. Calibration is performed with first priority against the time series of the number of hospital patients in Berlin, and with second priority against the COVID case numbers in Berlin. The case numbers are only used with second priority since the screening procedure has been changed multiple times, which means that the resulting time series is not homogeneous and thus not useful for model calibration.

The calibration includes the following elements:

1. Calibration of the basic doubling time without reduction of activity participation
2. Integration of spring disease import
3. Calibration of the consequences of reduced activity participation
4. Calibration of an indoors/outdoors effect for leisure activities depending on the temperature
5. Integration of contact tracing, masks, and summer disease import

All calibrations concern Θ (cf. Eq. 1); item 4 also involves defining threshold temperatures at which activities are moved outdoors at the end of the winter, and indoors at the end of the summer. All other aspects are data driven.

The final model is shown in Fig. 4 (top), where the blue line traces the number of new cases with state *showingSymptoms* from our simulation. Fig. 4 (bottom) shows the cases in need of hospital care and those in need of ICU care from our simulation compared to real data. As stated, we find fitting to the hospital numbers more important; fully fitting to the case numbers at the same time is not possible.

**Fig 4.**
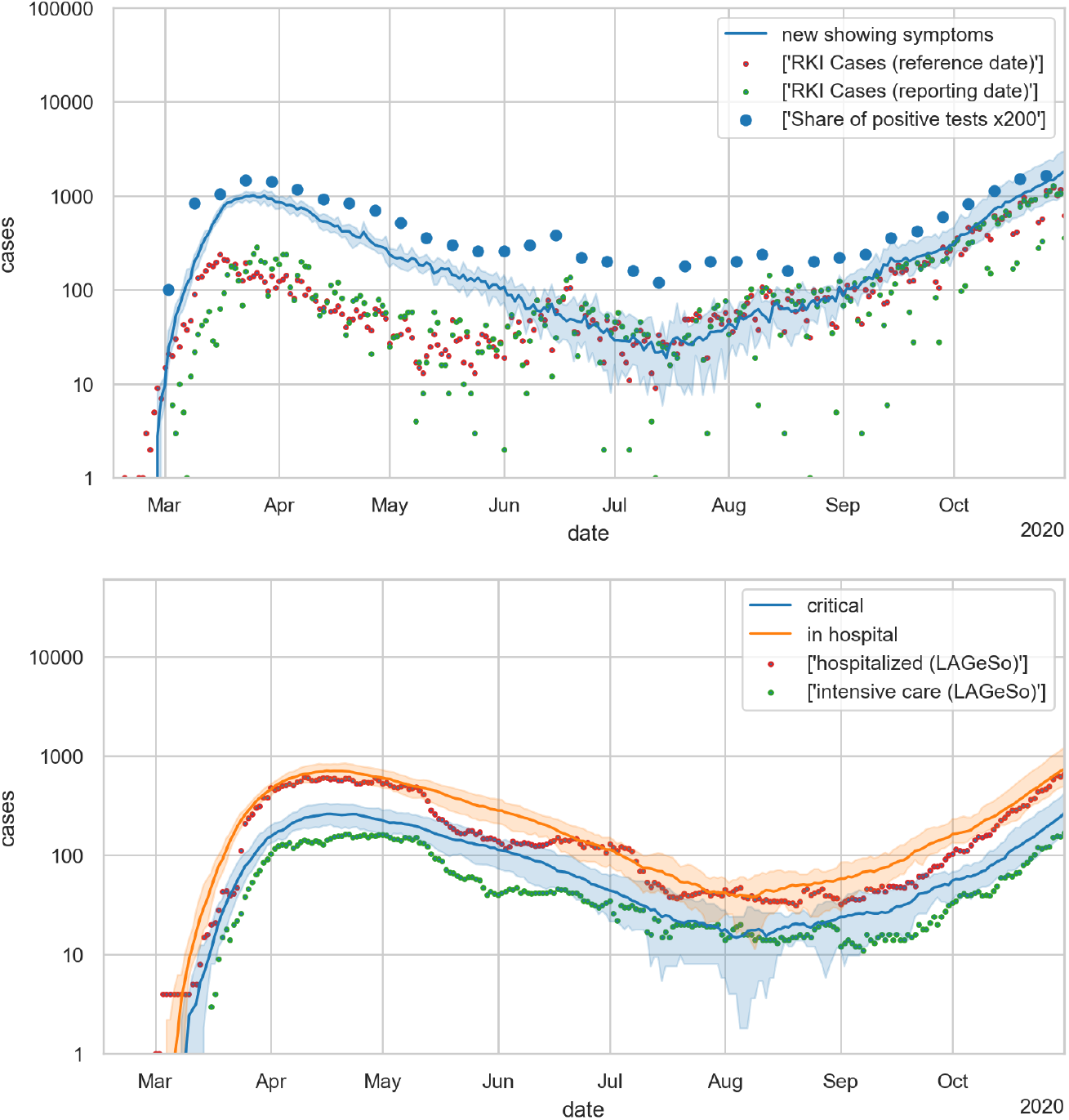
Final model. TOP: case numbers; BOTTOM: hospital numbers. https://covid-sim.info/2021-02-09/paperAggr?run=6_withSummerImport&withTracing=yes&withMasks=yes&thetaFactor=0.8&withMasksAndTracing=yes

**Fig 5.**
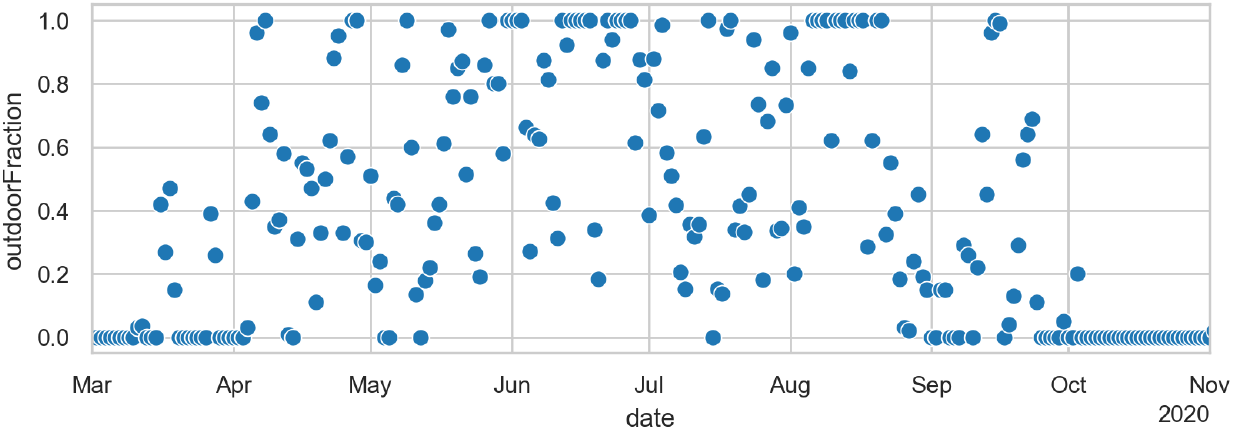
Outdoors fraction for activities of type leisure, depending on the temperature of each day.

In terms of calibration, the initial growth is, within limits, insensitive against changes of Θ, since it is dominated by the disease import (cf. Sec. Appendix: Calibration). This can be explained by the fact that the exponential growth was running ahead in other areas, and in consequence the *share* of infected persons from those areas also grew exponentially. Only after travel was stopped, disease import also stopped, and the dynamics in Berlin was dominated by internal processes. What *is* sensitive against Θ is the downward slope around April. In consequence, we adjust Θ such that the downward slope in the logarithmic plot is reproduced, given the activity reductions and mask compliance provided by our data. The result is also compared against hospital numbers (Fig. 4 bottom), which confirms our calibration.

The case numbers over the summer contain one large outbreak in a religious community, which the model does not contain. Otherwise, over the summer, the reinfection rate *R* was below one (Fig. 7).

**Fig 6.**
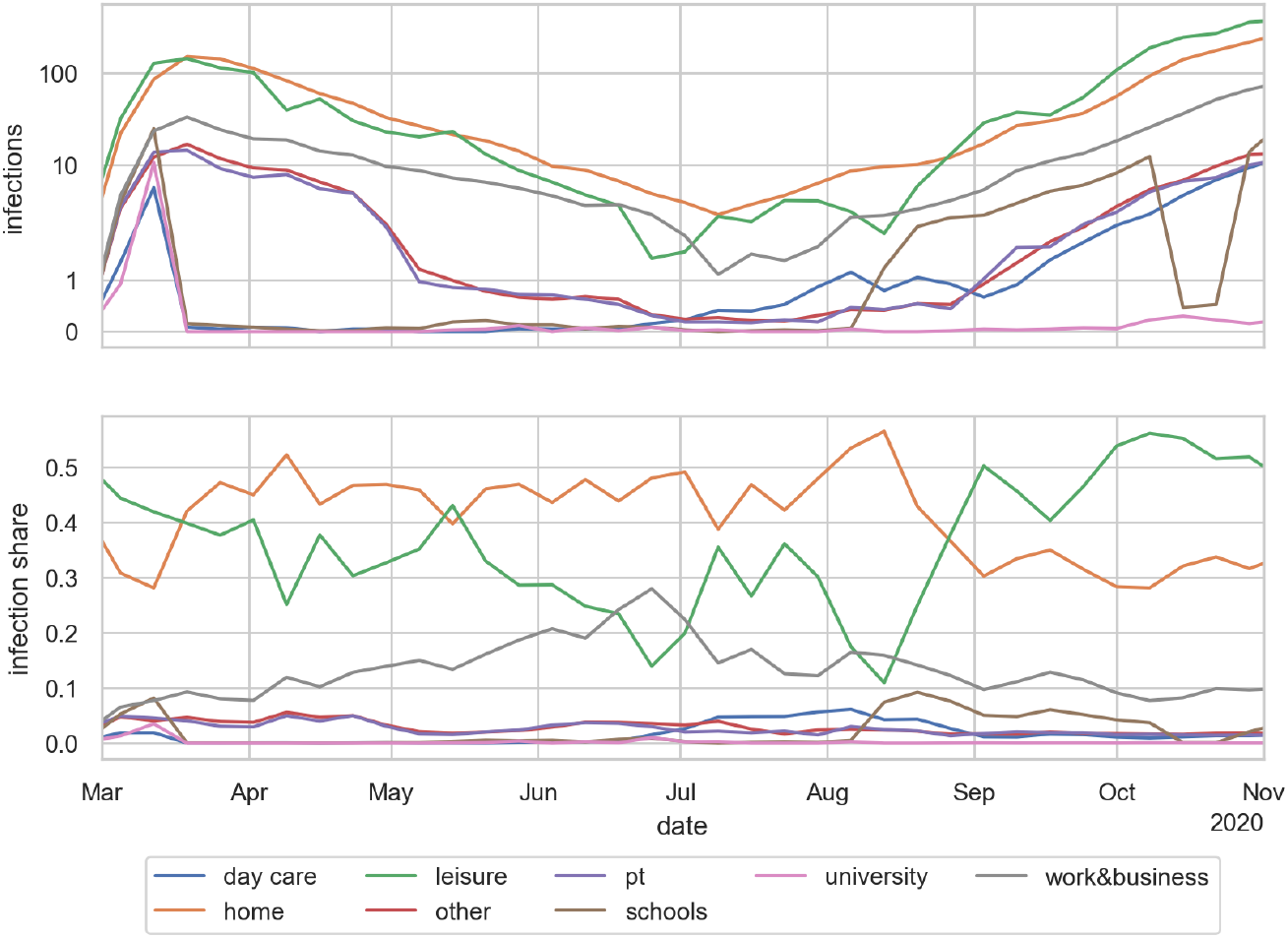
TOP: Infections per activity type. Note logarithmic scale. BOTTOM: Share of infections per activity type. The values are averaged over the same 10 runs as for the other figures, and in addition aggregated into weekly bins. One can see, for example, the return to school near the beginning of August, and the fall vacations in October.

**Fig 7.**
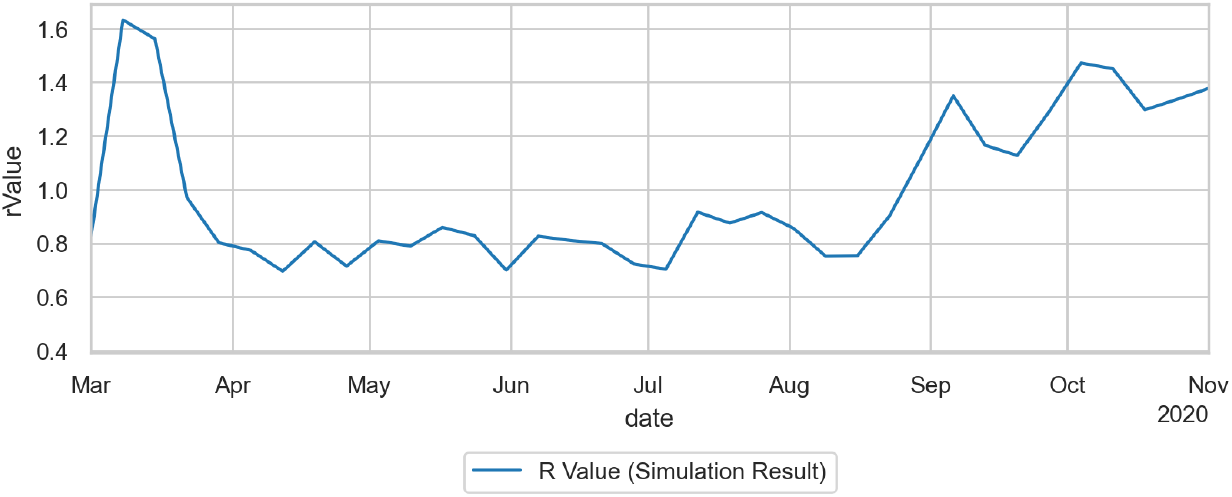
Reinfection rate *R*(*t*) for the duration of the simulation. As explained in the text, we explicitly count the reinfections per agent, and then average them over all agents that turned contagious on a given day.

### Indoors/outdoors and second wave

We include into our model that up to 100% of leisure activities are undertaken outdoors during summer, while that share reduces to 0% during winter. When an activity occurs outdoors, the otherwise identical computation of the infection probability is divided by 10. The model takes the actual temperatures as input; if the daily maximum temperature is larger than *T* ^*\**^ + 5*C*, then all activities that can happen outdoors are outdoors; if the daily maximum temperature is smaller than *T* ^*\**^*−* 5*C*, then all activities happen indoors; in between, probabilities are linearly interpolated. We use *T* ^*\**^ = 17.5*C* in spring, linearly increasing to *T* ^*\**^ = 25*C* in fall. Having different *T* ^*\**^ in spring vs fall is plausible, and yields a far more plausible infection dynamics than keeping them the same. More details are given in Sec. Outdoors vs. indoors season in the appendix.

The second wave, as one can see in Fig. 4, already started in July, in our simulations pushed first by disease imports from people returning from summer travel (cf. Fig. 10), and then by the school openings in the second week of August (cf., e.g., Fig. 8). However, our simulations say that, without the change in outdoor fractions, contact tracing would have been able to keep the dynamics under control after the returns from summer travel stopped. Also, models where the second wave is triggered by summer disease import plus school openings alone all result in a second wave that is “too early”, and also “not steep enough” when compared with data, in particular with hospital numbers. Weather is the only effect that we have identified so far that occurs at the right time.

**Fig 8.**
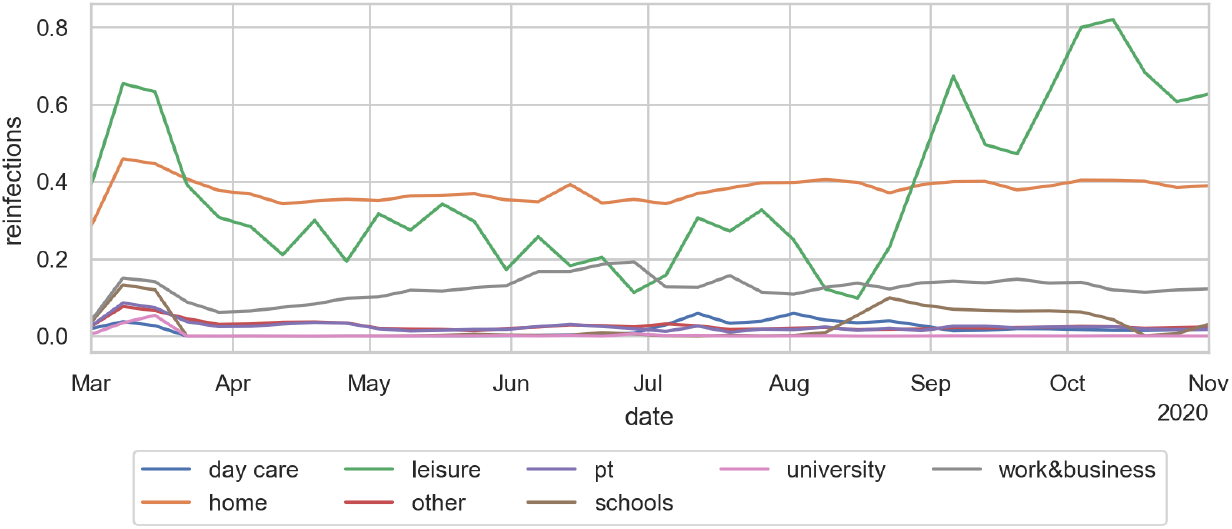
Reinfections per activity type.

**Fig 9.**
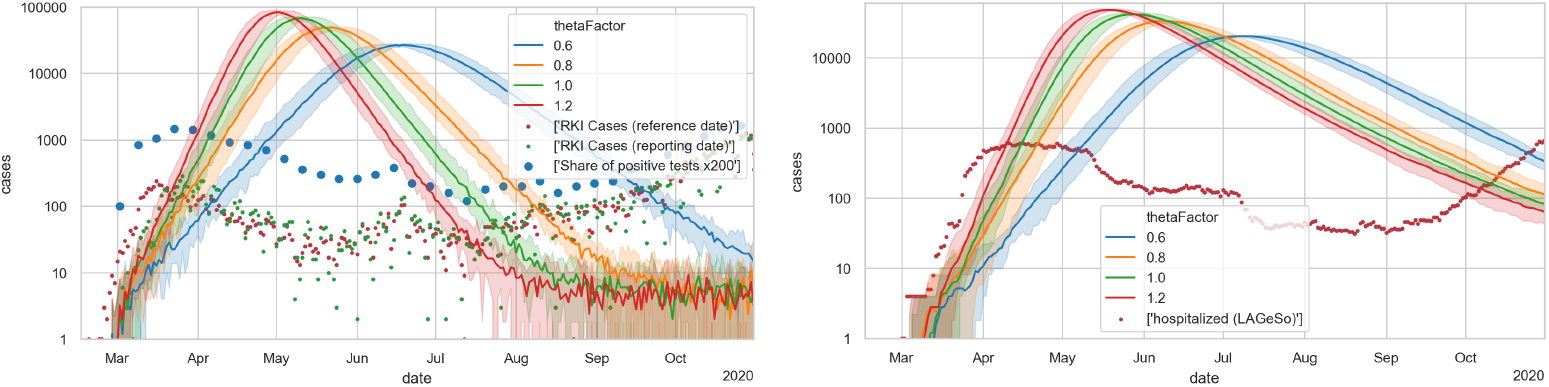
Unrestricted base case. LEFT: Case numbers. The green and red dots denote case numbers as reported by Robert Koch Institute [105]; the blue dots denote positive test fractions [106] multiplied by 200. RIGHT: Hospital numbers. Each simulation curve is averaged over 10 independent Monte Carlo runs with different random seeds.

**Fig 10.**
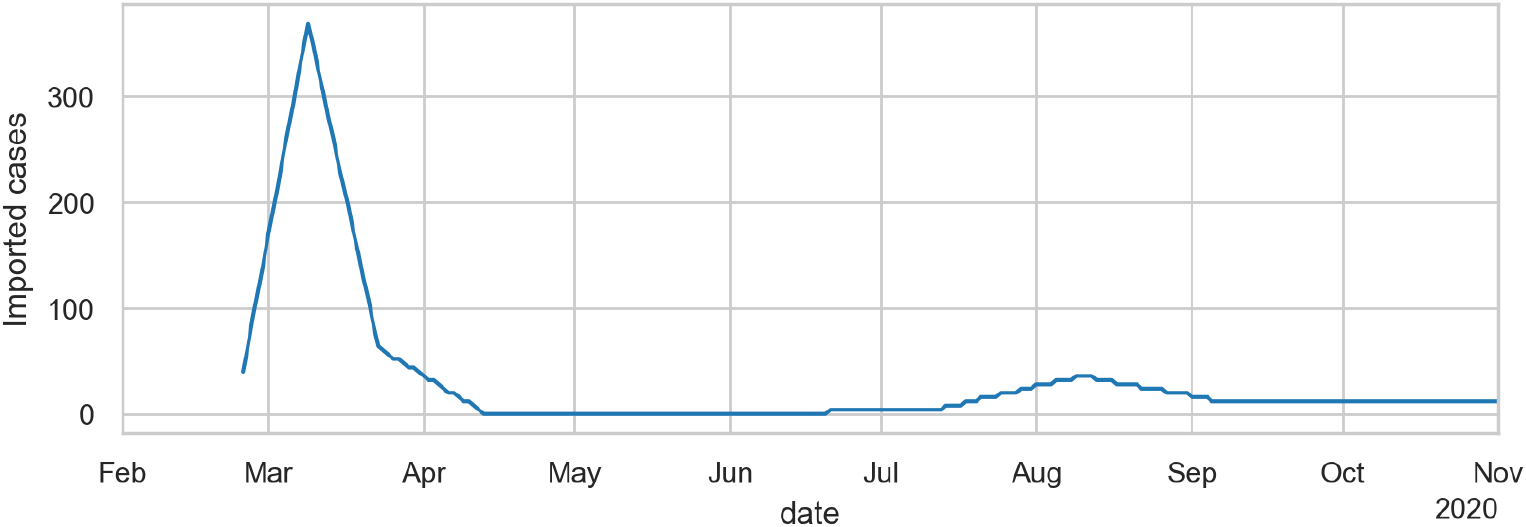
Disease import over time. Based on data taken from [105] (always on Tuesdays), but multiplied by 4 in spring, and divided by 2 in summer (see text for discussion).

### Infections per activity type

Evidently, in our microscopic models we can track how many infections happen at which activity type. Fig. 6 shows, on top, the absolute numbers of infections per activity type for the simulation, and below the *share* of infections per activity type over time. To obtain these numbers, we evaluate what activity the infected person is performing at the time of infection and date that to the date of infection.

Initially, all activity types play a role. After the closure of the universities, schools, and day care in March, both their absolute numbers and their shares go to zero. At the same time, the infections share of work (gray) in April and May reflects that persons were drifting back to normal activity patterns (cf. Fig. 2). Leisure (green) would have shown the same trend, but that was counter-acted by the increasing shift of activities to outdoors. In the bottom plot, the purple line shows how the share of infections in public transit decreases significantly near the end of April because of increased wearing of masks. (Recall that we use observed mask compliance.) In July we see how day care (blue) picks up, because it was re-opened. Schools re-open in the second week of August, and pick up accordingly (brown). Also, two weeks of school vacation in October are clearly reflected in the brown curve. From September on we then see a strong increase of the infections share of leisure activities – corresponding to moving leisure activities from outdoors to indoors as explained earlier.

### Reinfections

Since our method is person-centric, we can, for each infected person *n*, count the number of its reinfections, *R*_*n*_. When averaging over multiple persons, one needs to make a decision to which date *R*_*n*_ is assigned. We use the date when *n* turned contagious, and in consequence

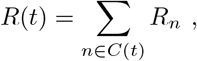

where *C*(*t*) refers to all persons who turned contagious on day *t*. An issue with this approach is that the consequences of interventions become visible in *R*(*t*) before the interventions actually start – since the reinfections that are suppressed happen later than *t*. This is also the reason why we use the date when turning contagious and not the date when they got infected, since that would increase that distance even more. Fig. 7 shows the resulting values, with *R*(*t*) much larger than one in the initial phase, then lower than one until the end of summer, and then increasing to above one in fall. We do not offer a comparison with the official R values since they have the same issues as the official case numbers.

### Reinfections per activity type

More insightful than the number or share of infections, as discussed in Sec. Infections per activity type below, are the average *reinfections* in each activity type. The method counts for each infected person the number of persons they reinfect at each activity context. As in Sec. Reinfections above, the numbers are dated back to the date when the person became contagious, and then averaged over all those persons.

One sees, in Fig. 8, that the reinfections at home remain roughly constant – a person who gets infected in any way in the average reinfects about 0.35 persons at home. Work is related to the mobility data – if less time is spent out-of-home, then in the model less time is spent at work, leading to fewer infections. Schools were closed in the middle of March, and not reopened until the second week of August. Also, there is a vacation during the second and third week of October. Day care according to the model has little effect. Day care was already re-opened partially in June, and fully in July. The reinfections at leisure are strongly driven by the weather: If it is warm, the model assumes that most of them take place outdoors, where they contribute little to the infection dynamics. In consequence, this effect plays an important role in spring, where the warmer temperature played as much a role as the reduction of the out-of-home activites. One also clearly sees the strong growth of the leisure reinfections in fall, which according to these simulations is driving the second wave in Berlin. Public transport is strongly visible in March, until the obligation to wear masks was introduced. All other infection contexts, e.g. errands or business activities, are combined in the category “other”.

### Reductions of R per intervention

Other papers, e.g. [12, 87], report, for various interventions, corresponding percent reductions of *R*. Our model clarifies that it is structurally more robust to report the *additive* reduction of the reinfections by a certain intervention. For example, according to our model closing schools removes the school reinfections from the dynamics, and in consequence reduces *R* by about 0.15. If *R* is 1 when the intervention is introduced, this amounts to 15%; if *R* is 2, then this amounts to 7.5%.

Tab. 2 shows, based on simulations as explained in the previous section, the contributions to R of the different activity types. Adding up the boldface numbers leads to *R* = 2.26, i.e. a strongly super-critical situation. In contrast, the 2020 Germany summer regime corresponds to closed universities, schools and day care, and wearing masks in retail. Together with the leisure summer number this leads to *R* = 0.88, i.e. makes the situation sub-critical.

**Table 2.**
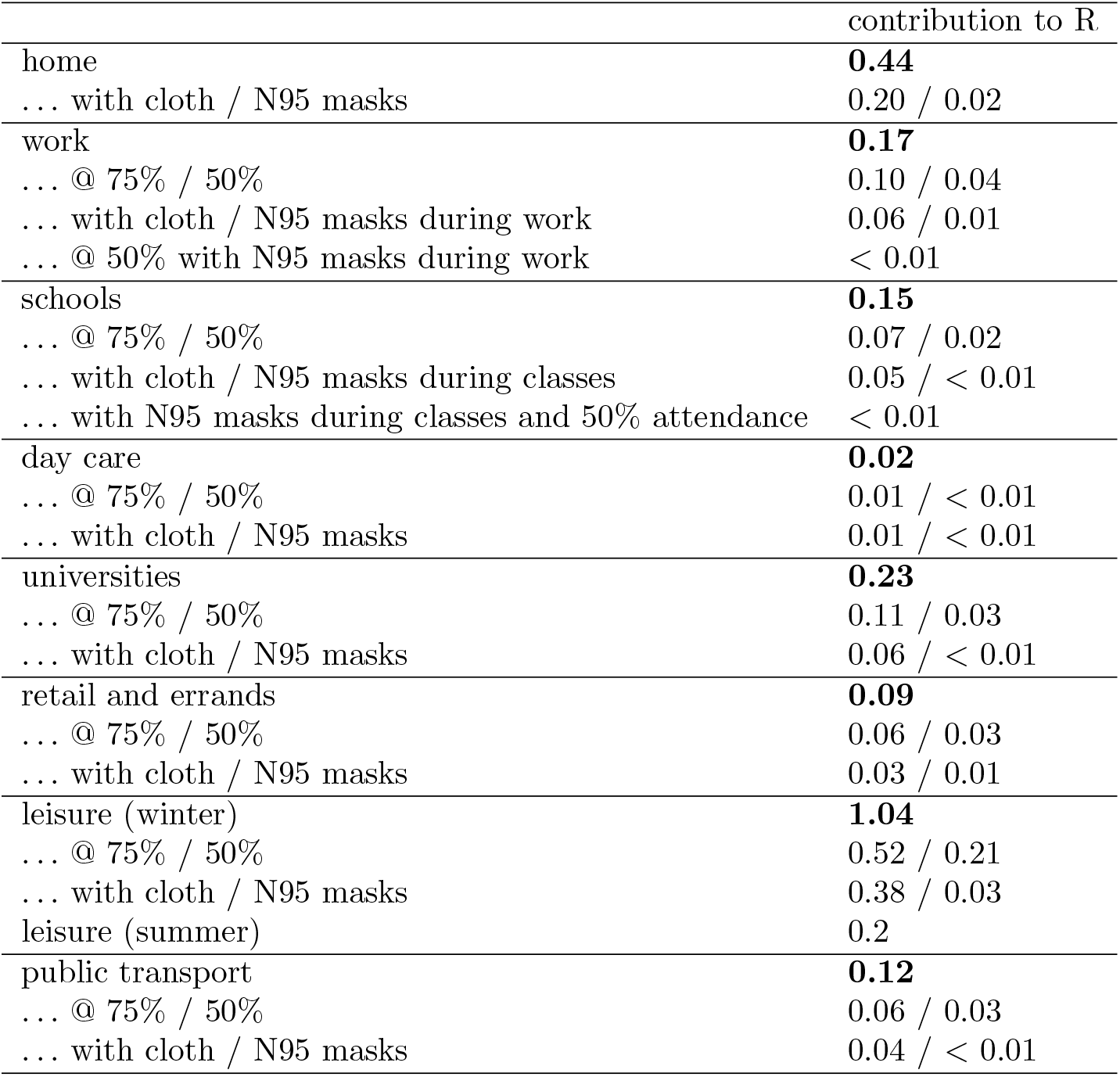
Contributions to R by activity type and intervention according to our model.

It has been pointed out by other studies that the reinfections at home play an important role and reduce the remaining “space” one has available for infections outside home [88]. Reinfections at home can be reduced by moving persons showing symptoms, and more radically persons identified as contacts by contact tracing, into separate facilities, sometimes called quarantine hotels.

One also notices that all infection contexts can be strongly reduced by wearing masks – this (evidently) even holds for leisure. Clearly, they would need to be worn *during* the activities, and not just during access and egress. Wearing masks during class at school has hesitantly been adopted in Berlin during November; wearing masks during work, in particular in office buildings, has never been pursued seriously in Germany and is still not obligatory if occupants have at least 10 *m*^2^ available per person – which is the value with which our simulations run and which generate the numbers of Table 2.

Evidently, a tricky context is leisure. According to our simulations, leisure alone, in conjunction with home, would be sufficient to keep R above one during winter, and thus needs to be suppressed accordingly. Keeping other activity contexts open without masks implies that leisure needs to be suppressed even further if *R <* 1 is to be achieved.

Conversely, during summer achieving an *R <* 1 is relatively easy. This explains why there were few problems during summer in Germany (and most other European countries).

### Decreasing marginal effect of interventions

Our approach clarifies why the marginal effect of stay-at-home interventions decreases [13]: Assume, for example, that each morning each school child throws a coin and goes to school only when it shows heads; this means that school participation is reduced by 50%. In consequence, if there is an infectious person at school, only half as many other persons have a chance to get infected. (Note that this assumes that they use the same classrooms as before, at half the density.) However, the probability that an undetected infectious person goes to school is also reduced to 50%. Multiplying these two probabilities means that only 50% *·* 50% = 25% of the infections happen in this case. More generally, reductions of activity participation have quadratic effect; a reduction of the participation to *α* leads to a reduction of infections to *α*^2^. Evidently, this means that 1 *—*0.75 *·* 0.75 = 44% of the effect is obtained with the first 25% of the intervention, 1 *—* 0.5 · 0.5 = 75% of the effect are obtained with the next 50% of the intervention, and the remaining 25% of the effect need the remaining 50% of the stay-at-home intervention.

In terms of the management of COVID-19, this implies that it is far better to include each activity type/sector of the economy to some extent, rather than shutting down some sectors completely while leaving some other sectors completely open.

### Intuition for these results

In an older version of the model [89], we had all contact intensities set to one. The contributions of each activity type to the infection dynamics then in first order corresponded to the average weekly time consumption in the respective activity. For example, averaged over the week including the weekend, school consumes about 5 hours per day for persons going to school. However, since in Berlin only about 10% of the population are school children,^1^ the average time consumption for the school activity is only 0.5 hours per day when taken across the whole population. In contrast, there are more persons going to work than to school, thus increasing the weight of work in the infection dynamics. The by far largest weight, however, comes from the leisure activities, which are not necessarily more hours per week for each individual person, but where *all* persons contribute to this type of time consumption. In consequence, restricting leisure activities had a large effect in that model.

In the present model, the time consumptions are now divided by the air volume flows per person in those activity types, cf. Tab. 1. In consequence, leisure, which already had a large share before, is now divided by a small air volume flow per person, and in consequence now gets even more weight. Work, despite occupying similar amounts of time, is weighted down because of the division by the much larger air volume flow per person. On the other end of the scale, public transport has, at full occupancy, a small air volume flow per person, but the times spent in public transport are considerably smaller than, say, at work. Also, persons in public transport are required to wear masks, while at work they are not.

A complicated case are schools and day care: They occupy large amounts of time, *and* have a small air volume flow per person, both somewhat similar to leisure. In consequence, the re-opening of day care in July and of the schools in August should have had strong consequences in the infection numbers. We took the observation that that did not happen as confirmation that their larger-than-average contact intensity is compensated for by a smaller-than-average infectivity and susceptibility. Clearly, this is specific to (our current understanding of) COVID-19.

For other diseases, for example influenza, all of the above may need to be adapted. For example, children may have a larger infectivity/susceptibility than adults, which then multiplied with their large contact intensity would lead to a large contribution to the infection dynamics. In consequence, these sub-models need to be understood and re-calibrated for each individual communicable disease.

## Discussion

### Comparison to compartmental models

Arguably, compartmental models are the mainstay of epidemiological modelling. Our approach, in contrast, follows individual synthetic persons. These individual persons can be enriched by person-centric attributes such as age or individual risk factors. Disease progression is individual, taking into account these demographic and other person-centric attributes. Similar to compartmental models, the base reinfection rate and the starting date need to be calibrated from case numbers (for the present study, the latter was replaced by data-driven disease import). However, both the spatial and the social interactions in our model come directly from data. Also, behavioral reductions in activity participation come directly from data. Mechanical aspects such as the wearing of masks by certain persons and/or at certain activity types can be integrated very simply into the model, by reducing virus shedding, virus intake, or both. Travel in public transport is already integrated. Organizational suppression approaches, such as contact tracing, can be simulated mechanically, thus extracting information about the allowed delays between symptom onset and reaching contacts, the failure rate, etc.

We were able to bring this up quickly: Coding of the infection code was started at the end of Feb/2020; our first preprint is from 20/Mar/2020 [90]; our first report to the government is from 8/Apr/2020 [10]; we have reported to the government regularly since then.^2^ Evidently, we were drawing from our experience and expertise with person-centric travel models. Still, it means that given the right experience and data availability, the method is not overly heavyweight, and then has many advantages over compartmental models.

The basic behavior of the model is like that of any S(E)IR model, i.e. exponential growth until a sufficient share of the population is immune, followed by exponential decline (cf. blue line in Fig. 4). Also the beginning and the speed of the growth are calibrated in similar ways. In typical S(E)IR models, however, interventions such as reductions in out-of-home activity participation, masks, or contact tracing, need to be parametrized into parameter changes of the S(E)IR model, most notably the infection rate [91–94]; in our model, such interventions are included directly into the corresponding processes.

A model that is at the border between compartmental and agent-based is by Chang et al. [40]. Important differences to our model include:

- Chang et al. take their movement model directly from mobile phone data. We, in contrast, re-use a pre-existing, activity-based model from transportation planning. This may be an advantage in regions where such a model already exists, and in particular so if the differentiated data that Chang et al. have is not available (as seems to be the case in Germany).
- We can attach individual attributes to each agent. In the present paper, this is used to model age dependence, a future study will contain the virus mutations, but it could also be used to include, say, pre-existing conditions. Compartmental models can only achieve this by introducing partial densities in each department. They will need as many partial densities as there are attribute combinations, i.e. *N*_*ageGroups*_ *· N*_*mutations*_ for the above situation. Since this is also multiplied with the number of locations, such models eventually need (much) more memory than when the information is attached directly to the individual agents.
- Both models are similar in that the conditional infection probability given contact is indirectly proportional to the floor area, for us stated in Eq. (3), and for Chang et al. stated in Eq. (8) in the appendix. We take floor area from maximum occupancy (obtained from the person trajectories) plus the typical surface area per person for each activity type. Chang et al, in contrast, take the floor area directly from their POI data. However, they do not explicitly consider workplaces or schools, and rather replace this by a parameterized “localized infection”; here, our model is more specific and thus more conducive to the consideration of explicit measures such as the introduction of masks, reduced activity participation, etc. Our model also takes typical air exchange rates into account.
- Our model, since it comes from transport planning, includes encounters in vehicles of public transport.
- Another difference, which, however, has nothing to do with the methodology, is that we calibrate and validate against hospital cases. This provides, at least in Germany, a more stable basis than case numbers, since in Germany the sampling strategy for PCR testing has changed several times, thus making the case numbers problematic as a time series.

### Comparison to other agent-based models driven by mobile phone data

As stated earlier, an approach similar to ours is by Aleta et al. [58]. There are the following differences:

- Aleta at al, similar to Chang et al, take their movement model directly from the mobile phone data. We, in contrast, use a pre-existing model from transportation planning. This is useful in particular in places where such a model already exists.
- Importantly, we also use the reduction-of-mobility data as input to our model.
- Aleta at al. use a model of 2% of the real population (85,000 synthetic persons) while ours consists 25% (1.25 mio). With models sampled at fraction *α*, one needs to make a decision if either one synthetic person stands for 1*/α* real persons, or if one models a fraction of the real population. We found the second path more intuitive. However, one needs to make sure that household sizes, group sizes at offices, etc., remain realistic. For household sizes, this can be achieved by synthetically constructing them, as both Aleta et al. and we do in some way. For all other locations, one needs to aggregate 1*/α* locations of the same type into one location in order to have realistic contact probabilities. Aleta at al. state that because of the sampled population, “colocation events between individuals are still quite sparse”, which points exactly to that issue.
- One issue with small population samples is that this makes the introduction of different virus strains difficult: One can essentially only introduce them in packets of 1*/α*, missing the relatively long early phase where their numbers are still low. We will report on this in a future paper.
- In contrast to Aleta et al, we use an infection model constructed from first principles. This would presumably be easy to change in their model.
- Where we use lognormal distributions, Aleta et al. use exponential distributions (i.e. rates) to transition from one disease state to the next; we believe that the literature prefers lognormal distributions. Using exponential distributions will lead to wider distributions of disease durations, both with shorter and longer durations from infection to recovery. This also would presumably be easy to change in their model.

### Comparison to other “reductions of *R*” studies

Tab. 3 is an attempt to extract “additional reductions to *R*” from other studies. One immediately finds two issues: (A) The categories are not well aligned. For example, “small gathering cancellation” refers to gatherings with 50 persons or less, while other studies cancel gatherings *larger* than a certain number. Again other studies just consider a “gathering ban”, but at the same time have “event ban” and “venue closure” as separate items. (B) Even where the categories are well aligned, the resulting numbers vary widely: “schools closed” goes from 5% to 50%, “national lockdown” goes from 0 to 81%.

**Table 3.** Percent reduction of *R* in other studies. Rounded to integers.

| Measure | [87] | [95]<br>“CC” | [95]<br>“LASSO” | [96] | [97] |
| --- | --- | --- | --- | --- | --- |
| Schools closed | 50 | 16 | 34 | 8 | 5 |
| Some businesses suspended | 26 |  |  |  |  |
| Most businesses suspended | 34 |  |  |  |  |
| Work ban |  |  |  | 31 |  |
| Gatherings limited to $\leq 1000$ | 16 | | | | |
| Gatherings limited to $\leq 100$ | 17 | | | | |
| Gatherings limited to $\leq 10$ | 28 | | | | |
| Mass gathering cancellation ($> 50$) | | 27 | 0 | | |
| Small gathering cancellation ($\leq 50$) | | 17 | 23 | | |
| Event ban |  |  |  | 23 | 5 |
| Gathering ban |  |  |  | 34 |  |
| Venue closure |  |  |  | 36 |  |
| Stay-at-home order with exemptions | 14 |  |  |  |  |
| National lockdown |  | 0 | 8 | 5 | 81 |
| Mask-wearing | 0* |  |  |  |  |
| Increase availability of person protective equipment |  | 9 | 12 |  |  |

\* not significantly different from zero

In part, this is a consequence of the fact that the interventions are not standardized: For example, the number of exemptions in what is called a lockdown varied quite a lot between countries.

Additionally, the transmission mechanisms vary, so even if the concept may be the same, the execution and thus the effect may be quite different between countries. For example, our reductions to *R* caused by school closures come out much lower than most other values, in particular those of Brauner et al. [87]. We attribute this to the following two elements: First, the model by Brauner et al. has no initial disease import which is then brought to a halt. In consequence, their approach has to assign all changes in the infection dynamics to the school closures. The school closures in Berlin were, however, one week later, with Mar/12 (fri) or Mar/15 (mon) as the last day of school; were too late to explain the infection numbers. Also, Dehning et al. [91] have an additional change point on Mar/7, corroborating that something has changed before the school closures. Second, other than both Brauner et al. and Dehning et al., we have the mobility data of Fig. 2 at our disposal. It is clear that there was considerably more societal adaptation around the weekend of Mar/13-14 than just keeping children at home. Brauner et al. themselves write that “the closure of schools … may have caused … behaviour changes. We do not distinguish this indirect signalling effect from the direct effect”. Additionally, in Germany, children staying at home will force their parents to stay at home, thus forcing them into home office. In consequence, some of this may not be signalling, but causal secondary effects. In consequence, our model is more differentiated: What Brauner et al. attribute to the school closures alone is in our model attributed to a combination of school closures, behavioral changes, and the reduction of various other out-of-home activities. Thus, all of the values may be correct: The pure effect of school closures in western countries (with relatively few young people) may not be larger than 5%, but the measurable consequence for *R* when governments closed schools as their first intervention presumably indeed was much larger.

We have checked our relatively large reductions of *R* for masks multiple times. They are a consequence of the assumption that N95 masks reduce intake to 2.5%, taken from [98]. The review article [99] comes up with about 5%, a factor of two larger, but still displaying a very large reduction. The same paper [99] also shows that “masks” without a specification of the type has much less of an effect. Finally, there may be the issue that lay people may not be able to use N95 masks at full efficiency. In consequence, our results have to be interpreted once more “mechanically”: They are plausible under the assumption that the fraction of people specified in the model is indeed able to use N95 masks effectively.

Clearly, data-driven mechanical models such as ours help clarifying the categories since we can exactly specify what we mean by closing some activity type or wearing a mask at certain activity types. Also, we can differentiate between the transmission from political decision to behavioral execution vs. the consequences of the behavioral execution to the infection dynamics. Finally, we can mechanically include organizational approaches such as contact tracing.

### Under-reporting

A known issue with epidemiological data and thus the simulations that build on it is the issue of under-reporting, i.e. that there are more infections in reality than are in the data. Looking at Fig. 4, it is clear that our current model assumes only little under-reporting during August to October. This originally led to hospital numbers that were too large; since we cannot reduce the number of infections below the case numbers, this justifies why we reduce simulated hospital numbers by a factor of 2 as stated in Sec. Calibration. This, in turn, implies that, if we want to get the spring hospital numbers right, our simulated infection numbers in spring need to be about a factor of 8 larger than the case numbers. Also note that our simulation includes non-symptomatic cases, which come on top of the symptomatic cases that we show in our figures such as Fig. 4; that is, actual under-reporting is even larger. Still, it is entirely possible that the testing strategy is missing even more cases, in which case the simulation would need to aim for even larger numbers of infected persons. As long as the number of sero-positive persons in Germany remains in the single-digit percentage ranges [100], the predictions made by the simulation are not strongly affected by this issue. Once the infections start to saturate, i.e. approach herd immunity, this will become important. Hopefully, by then systematic antibody screenings will be available, and we will be able to calibrate the model against the case numbers that must have been infected in the past. Given that we have the hospital numbers for control, we expect this to be straightforward.

### Contact structures

It is well established that different contact structures lead to different infection dynamics [47, 101]. For example, the epidemic threshold, i.e. the minimal share of persons that need to be susceptible, may be different. For our model, such elements in principle come from the input data: Besides coming with their complete contact graph, our synthetic persons have an age, an employment status, and gender. Thus, in principle, we have the interaction structure at the level of facilities or public transport vehicles, including people’s attributes, from data. However, as stated, for privacy reasons the facilities are too large, and in consequence multiple households or multiple offices are bundled into a single facility. For households we compensate, as described, by manually splitting them up; as of now we do not, however, control for age structure in the splitting process. This needs to be improved. For all other activities we compensate, as also described, by allowing interaction with only 1*/N*^*spacesPerFacility*^ th of all other persons at the same facility on each given day, but allowing mixing by using separate random draws for every simulated day, selecting *N*^*spacesPerFacility*^ such that the overall number of contacts (for contact tracing) ends up in a plausible range. This should be improved as well. However, more of an issue for COVID-19 may be that the original input data does not contain separate facilities for the elderly; for mobility modelling, this has so far not been of interest. This implies that the following issues need to be addressed in the future:

- Clarify which epidemiology-relevant aspects of the input data are too far away from reality.
- For those aspects that need to be improved, clarify if this could be done at the level of the original input data generation, or if it should be compensated for at the level of the modelling.

Concerning the possibly different epidemic threshold, we would argue that for the present situation this is less of a problem: If only a fraction of the persons in the simulation was susceptible, or they would be connected via a different contact structure, the calibration process would compensate by selecting a different Θ in Eq. (1). Evidently, if we get closer to herd immunity, possibly by vaccination, these aspects become more important.

### Predictions

The model is used for predictions. We decided to not add them into the paper since any prediction we make now would be historical quickly. Our regular reports to the government, and thus our predictions, all have a DOI, for example [10] or [102].^3^

## Conclusions

We combine a person-centric human mobility model with a mechanical model of infection and a person-centric disease progression model into an epidemiological simulation model. Different from other models, we take the movements of the persons, including the intervening activities where they can interact with other people, directly from data. For privacy reasons, we rely on a process that takes the original mobile phone data, extracts statistical properties, and then synthesizes movement trajectories from the statistical properties; one could use the original mobile phone trajectories directly if they were available. The model is used to replay the epidemics in Berlin. It is shown that the second wave in Berlin can be modelled well with an explicit temperature dependency of the outdoors fraction for leisure activities. The model is then used to evaluate different intervention strategies, such as closing educational facilities, reducing other out-of-home activities, wearing masks, or contact tracing, and to determine differentiated percentage changes of the reinfection number *R* per intervention.

## Data Availability

All input data is available; URLs are in the paper.

## Appendix: Calibration

### Case numbers and hospital numbers in Berlin

The simulation is calibrated against the Berlin case numbers and the Berlin hospital numbers. COVID-19 is a notifiable disease, and the notifications are collected and published by the Robert Koch Institute (RKI) [103]. Each record contains at least two dates: The date when the record reaches the local health department (reporting date), and the date when symptoms started, called reference date.

In principle, the reference date would be easier to compare with our simulations, since it corresponds to the onset of our *showingSymptoms* state. Unfortunately, however, it is not clear how reliable that date is. The health department becomes aware of cases once they are tested positively. The positive test result becomes available about 2 days after the probe was taken. The health authorities thus have to connect a positive test with the person, and query the person about when symptoms started. Self-reported dates of symptoms onset are presumably rather unreliable, in part because of recall errors, in part because what a symptom is is not sharply defined. The reliability may be improved by using expert interviewers, but those may not always be available. In addition, when tests are taken from pre- or asymptomatic cases, a date of symptoms onset is not yet available, and for asymptomatic cases never will be. In such cases, the reporting data is also entered as reference date, which for pre-symptomatic cases is too early. Finally, many records are reported completely without this reference date. RKI provides a procedure to impute the missing reference date [104], but has to rely on the statistical distribution of the cases where a reference date exists, which may not be a valid assumption since, say, locations that are under stress of high infection numbers may both not enter the reference date *and* receive the test results with additional delay. Also, the sampling strategy for testing was changed several times.

In consequence, we plot the case numbers both by reporting and by reference date for comparison, and also add a third number: The fraction of positive tests. In a targeted testing regime, this fraction will go up when testing is made more restrictive, and the other way around. It will thus react to changes in the testing regime in the opposite direction as the case numbers. In practical terms, we normalize the fraction curve such that it coincides with the cases curve in fall, and is above the cases curve during all other times. This leads to a plausible corridor for the simulations.

Because of these issues, we also calibrate, in fact with higher priority, against the hospital and the ICU numbers in Berlin. We believe those to be relatively unbiased, since there was always sufficient hospital capacity in Berlin throughout the period considered here.

### Unrestricted model

Most parameters of the model are taken from the literature, as explained earlier, in particular Fig 3. The remaining free parameters are, from Eq. (1), Θ, *sh*, and *in*. We have set the base values of *sh* = *in* = 1. As mentioned before, we use these parameters to model the wearing of masks, meaning that they are reduced when masks are worn.

Fig. 9 shows the unrestricted base case with four different values of Θ. One finds that the aggregated behavior at this level corresponds to that of typical SE(I)R models, i.e. exponential growth, followed by a maximum, followed by exponential decrease. Based on these plots, thetaFactor values of 1.0 or 1.2 seem plausible to be consistent with the initial growth.^4^

### Spring disease import

We take the disease import from abroad from data published by RKI ([105], always on Tuesdays). Currently, for Germany this data is only available on a nationwide aggregated level. For this reason we scale it down to our Berlin model by using the population size. The data is dated on the reporting date and not on the actual date of becoming sick. Since the infection seeds are initiated into our model with the status *exposed* (cf. Fig. 3) and it can be assumed that the reporting date is significantly after the exposure date we date the data from RKI back by one week. The data provided by RKI is available as weekly values so we assign these values to the respective Monday and then interpolate between them. Since we assume unterreporting in the RKI numbers, we multiply them by 4; this is discussed in Sec. Under-reporting. The initially infected persons are drawn randomly from the population. The resulting disease import is shown in Fig. 10; the description so far only concerns the spring disease import.

An advantage about adding disease import is that the date of the first infection is no longer a free parameter: As shown in Fig. 11, the disease import is sufficient to drive the first wave. The disease import data seems to lack some early cases, thus causing an initially nearly vertical increase in the simulation. The dynamics then settles onto the exponential increase shown in the previous section.

**Fig 11.**
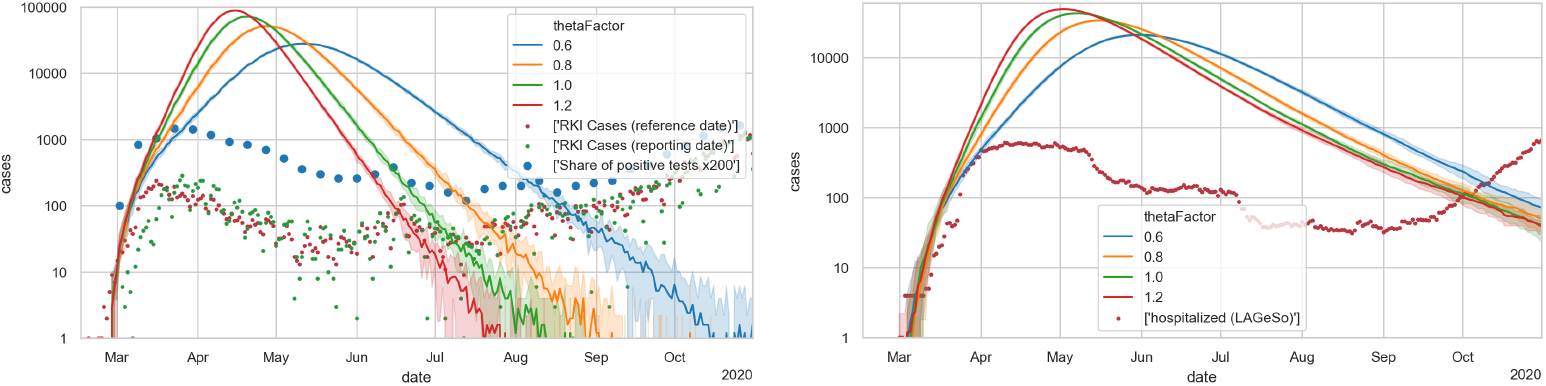
Unrestricted base case, but with initial disease import from data. LEFT: Case numbers; RIGHT: hospital cases. One finds that the initial slope dynamics is rather independent from the thetaFactor.

### Reductions of activity participation

During the unfolding of the epidemics, people decided or were ordered to no longer participate in certain activities. We model this by removing an activity from a person’s schedule, plus the travel to and from the activity. In consequence, that person no longer interacts with people at that activity location, and in consequence neither can infect other persons nor can become infected during that activity, or while in public transport vehicles to and from that activity. Overall, this reduces contact options, and thus reduces epidemic spread.

A very important consequence of our modelling approach is that we can take that reduction in activity participation from data. As stated earlier, that data comes from the same source as our original activity patterns. Unfortunately, the activity type detection algorithm is not very good for these unusual activity patterns, as one can see in Fig. 12 when knowing that all educational institutions were closed in Berlin after Mar/15. What is reliable, though, is the differentiation between at-home and out-of-home time, as displayed in Fig. 2. One clearly notices that out-of-home activities are somewhat reduced after Mar/8, and dramatically reduced soon after. After some experimentation, it was decided to take weekly averages of the activity non-participation, and use that uniformly across all activity types in our model, except for educational activities, which were taken as ordered by the government.

**Fig 12.**
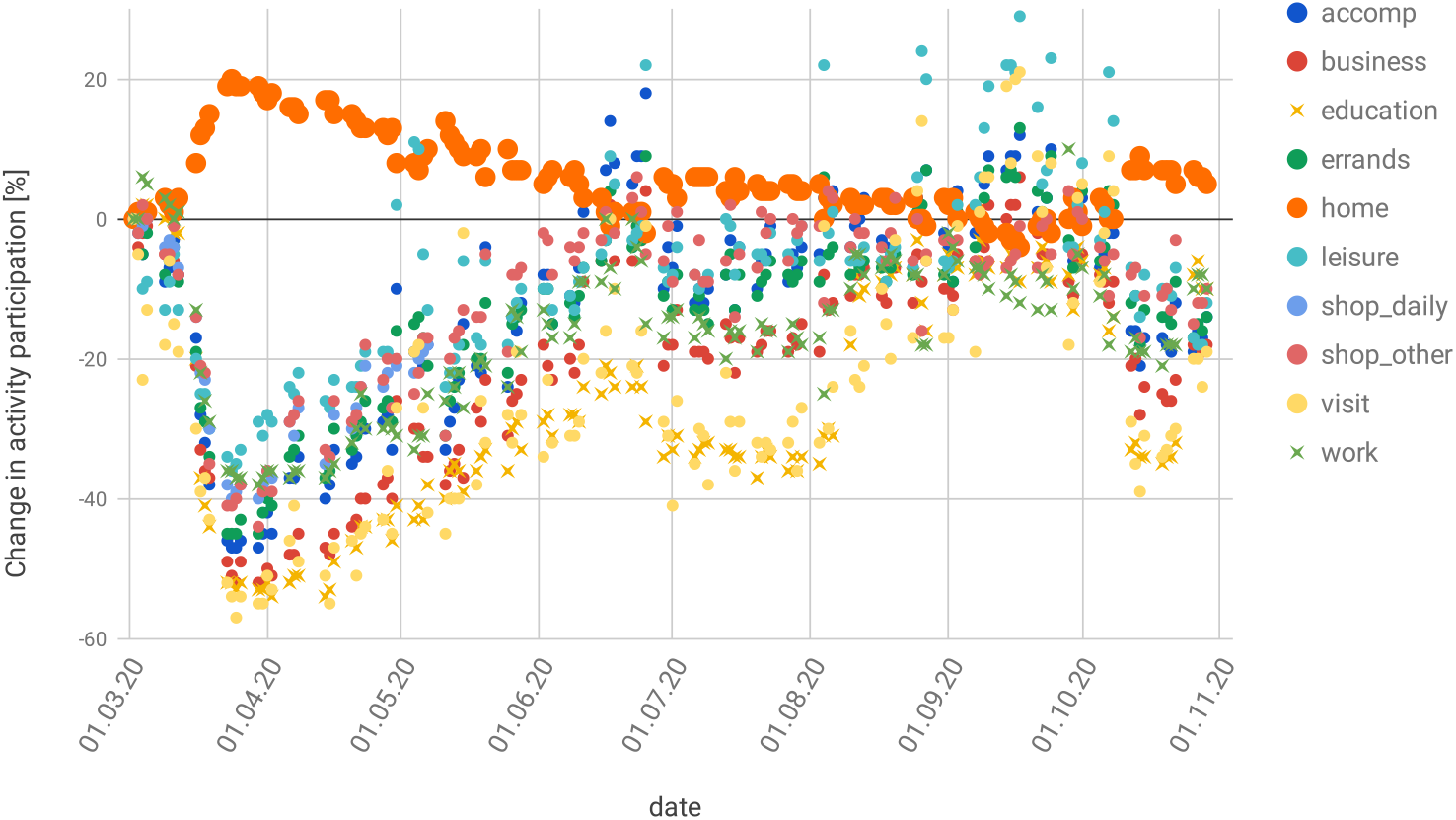
Reduced activity participation over the course of the epidemics in Berlin.

To remove an activity with a certain probability, a random draw is made every time a synthetic person has that activity type in its plan. This means that the model assumes that, say for a 50% work reduction, there will be another 50% subset of persons at work every day. This intervention, in consequence, does not sever infection networks, but just slows down the dynamics.

One takes from Fig. 13 that the mobility reductions, as given by the mobility data, is by itself not sufficient to explain the decreasing case numbers during spring. Evidently, one could now reduce Θ, and this is what we have done in our early simulations. This, however, artificially reduces the infection dynamics, and means that the simulation will miss the second wave in fall.

**Fig 13.**
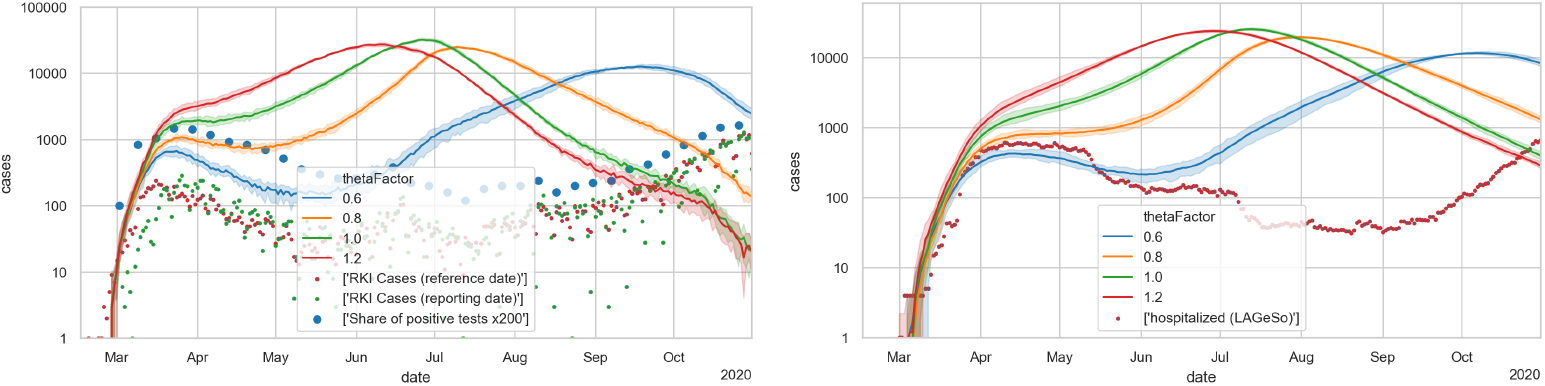
Case numbers (LEFT) and hospital numbers (RIGHT) in simulations with reductions of activity participation as obtained from mobility data.

### Outdoors vs. indoors season

The probability of getting infected during an encounter depends on whether the encounter takes place indoors or outdoors. Outside, the probability of infection is significantly reduced compared to inside. This is due to the fact that outdoors the air is constantly in motion and therefore aerosols cannot accumulate. We assume that an encounter outdoors decreases the infection probability by one magnitude [4, 9]. In countries like Germany, seasonality has a great influence on how much time people spend outside. In summer, people spend more time outdoors, while in winter they tend to spend more time indoors.

We include into our model that up to 100% of leisure activities are undertaken outdoors during summer, while that share reduces to 0% during winter. When an activity occurs outdoors, the otherwise identical computation of the infection probability is divided by 10. The model takes the actual temperatures as input; if the daily maximum temperature is larger than *T* ^*\**^ + 5*C*, then all activities that can happen outdoors are outdoors; if the daily maximum temperature is smaller than *T* ^*\**^*−* 5*C*, then all activities happen indoors; in between, probabilities are linearly interpolated. We use *T* ^*\**^ = 17.5*C* in spring, linearly increasing to *T* ^*\**^ = 25*C* in fall; runs with different *T* ^*\**^ in spring vs fall are plausible, and yields a far more plausible infection dynamics than keeping them the same.

The justification for this is as follows. A survey on physical activities [107] shows that, in summer, people in Germany perform about 80% of their physical activities outdoors, while this proportion shrinks to 10% in winter. We have assumed that other leisure activities (e.g. restaurants, visit friends) behave similarly. We also extend our range to 0 and 100% since the fluctuations of the temperature already lead to average values that are more than 0 and less than 100% (cf. Fig. 5).

Fig. 14 shows an example of the infection dynamics where both *T* ^*\**^ in spring and *T* ^*\**^ in fall are 17.5C; as one can see, either the decrease of the first wave is not strong enough, or the second wave comes too late; note in particular the hospital numbers, which for all values of thetaFactor do not have enough slope in the second wave. The results with other *T* ^*\**^, as long as they are the same in spring and fall, are the same. Fig. 15 shows using 17.5C for spring and 25C for fall; the second wave now is triggered earlier, and it is steeper.

**Fig 14.**
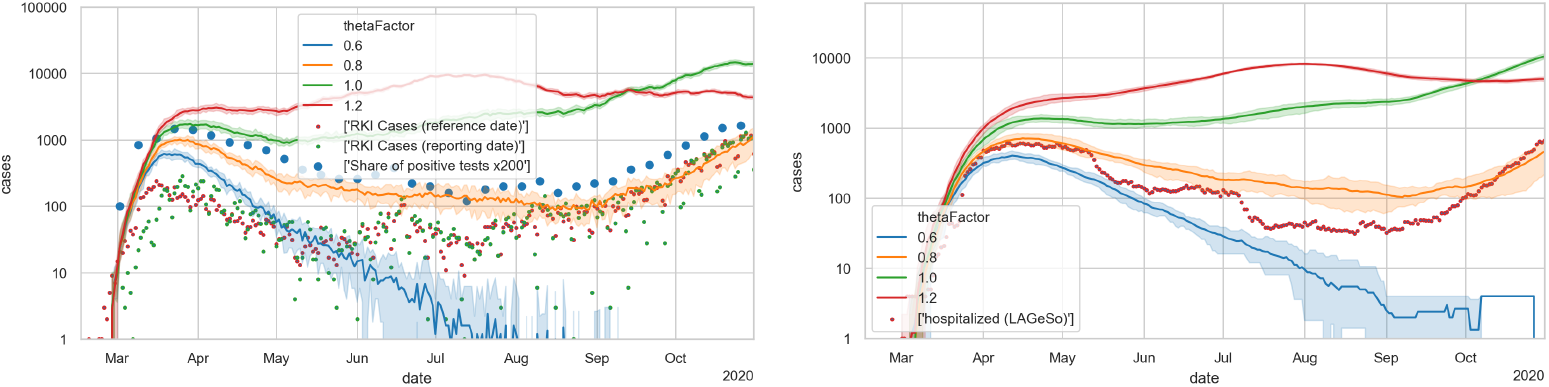
Case numbers (LEFT) and hospital numbers (RIGHT) in simulations that now also include the indoors/outdoors model, with a threshold temperature of 17.5C. A thetaFactor between 0.6 and 0.8 is most plausible, but the second wave would come too late and would not be steep enough (cf. in particular the hospital numbers).

**Fig 15.**
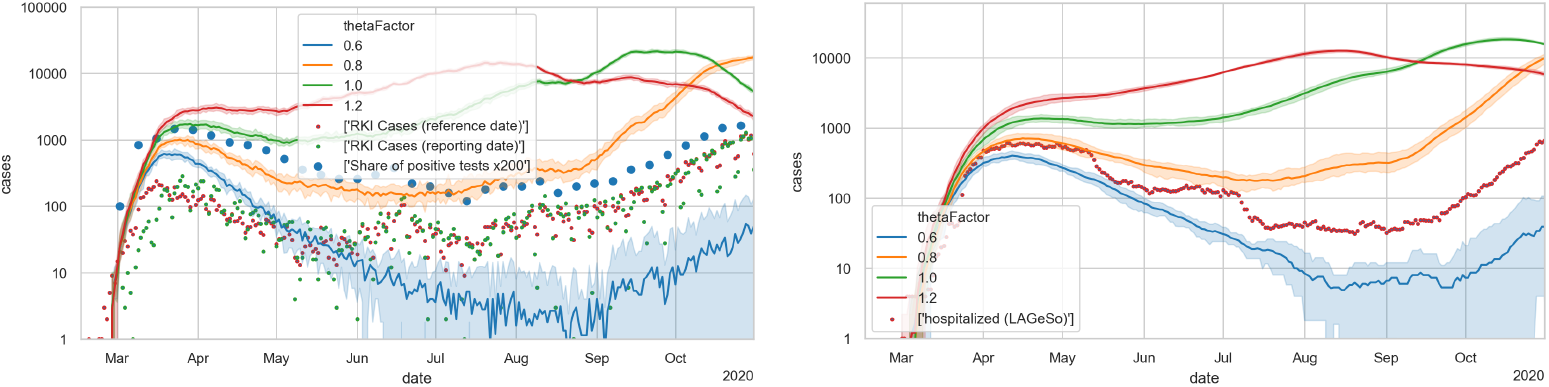
Case numbers (LEFT) and hospital numbers (RIGHT) in simulations that now also include the indoors/outdoors model, with a threshold temperature of 17.5C in spring, and 25C in fall. A thetaFactor between 0.6 and 0.8 is most plausible, which would well reproduce the second wave (cf. in particular the hospital numbers).

**Fig 16.**
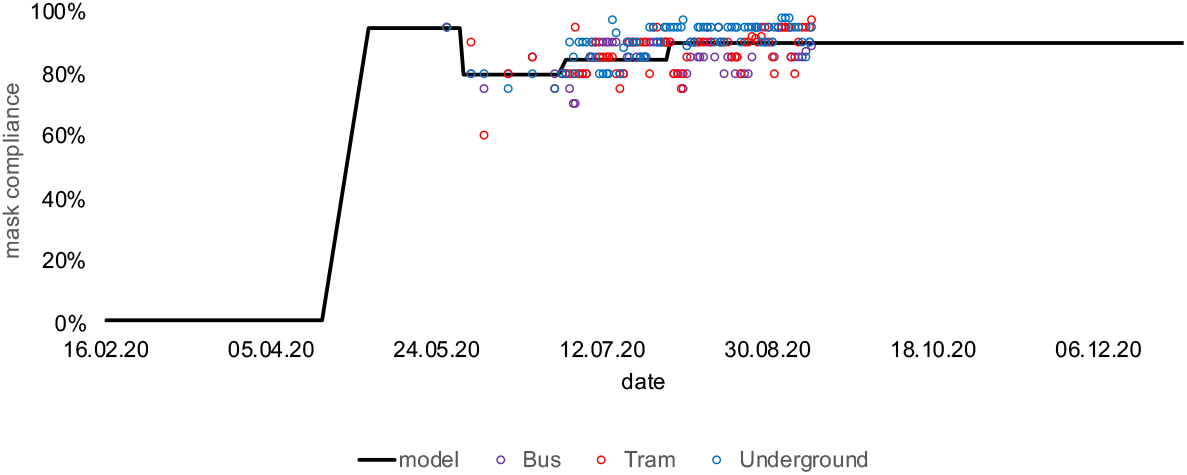
Mask compliance rates over time. Data provided by BVG [109].

### Masks and contact tracing

In April the wearing of masks in shops and in public transport vehicles became obligatory in Berlin [108]. We have included this into the infection model of Eq. 1 by reducing *sh* (if the contagious person wears a mask) and *in* (if the person to be potentially infected wears a mask). This is dependent on the activity type, meaning that persons only wear masks when shopping, doing errands or using public transport. The effectiveness of different mask types is taken from from [98], i.e. cloth masks reduce shedding and intake to 0.6 and 0.5 of their original values, surgical masks to 0.3 and 0.3, and N95 (FFP2) masks to 0.15 and 0.025. The review article [99] comes up with about for N95 masks, a factor of two larger, but still displaying a very large reduction. The same paper [99] also shows that “masks” without a specification of the type has much less of an effect. Finally, there may be the issue that lay people may not be able to use N95 masks at full efficiency. In consequence, any of our results that depend on N95 mask efficiency have to be interpreted “mechanically”: They are plausible under the assumption that the fraction of people specified in the model is indeed able to use N95 masks effectively.

The local transport company in Berlin (BVG, [109]) have provided us with the compliance rates in public transport over time meaning that we do not have to estimate them. We assume that the same compliance rates also apply to shopping activities. We assume that 90% of those people wearing masks wear cloth masks and 10% wear N95 masks.

The goal of contact tracing is to break chains of transmission by tracing the contacts of an infected person and putting these contacts into quarantine. In our model contacts are traced during all activities except for public transport and shopping because we assume that the health authorities are not able to find these contacts. A contact person is only traced when the contact duration is longer than 15 minutes, which corresponds to the RKI guidelines [110].

Persons that go into *showingSymptoms* are assumed to trigger a contact tracing mechanism, which works as follows:

1. Look at all traced contacts that the infected person had in the 2 days [110] before showing symptoms.
2. A probability *γ* determines if a contact person can be reached successfully and also follows the stay-at-home order. *γ* is set to 0.5
3. The persons that have been traced successfully go into quarantine, but only after a delay of *d* days, which allows to model the response time of the system. Our base value of *d* is set to 5 days. Personal experience in our surroundings says that tests are normally taken a day after symptoms start, and the result is available again one day later in the evening. That is, contact tracing can start no earlier than 3 days after symptoms onset. We add another two days to account for possible additional delays.
4. A tracing capacity limits the number of persons per day for which its contacts can be traced. The capacity is set to 0 until the end of March, 40 cases per day until 14/Jun, and to 200 cases per day afterwards. Germany had agreed on a limit of 50 cases per 100 000 inhabitants per week at which local governments were expected to act [111]. This number was based on what the system presumably could handle for contact tracing. For our Berlin scenario with 5 million persons, this translates to 357 cases per day. Based on newspaper reports [112], the system was overwhelmed already at lower numbers, which is why we use 200.
5. Persons leave the home quarantine after 14 days, if they did not develop symptoms during that time.

For *d*, a smaller value would be much better in terms of effectiveness, but our personal experience in several cases says that this is unrealistic. For *γ* and the maximum tracing capacity, we compared simulation results.^5^ Changes in *γ* make relatively little difference. For the maximum tracing capacity, one can see that larger capacities would have kept the new infections under control for longer than what happened in reality.

Masks and contact tracing, as described above, do not have a strong enough effect to gauge them from the infection or hospital numbers. As explained in Sec. Reinfections per activity type, masks in public transport and while shopping reduces R by about 0.1 each. Since masks were introduced in April, they reduce the slopes of all curves of Fig. 15 accordingly. This makes the blue curve from that figure less plausible and the orange curve more plausible, which is welcome since the larger thetaFactor is more plausible (cf. Sec. Unrestricted model).

Contact tracing, in contrast, just pulls the infection numbers down while they were low. Once contact tracing is overrun, it no longer influences exponential growth, and thus not the slopes of the second wave in the logplot.

We leave both of these elements in the model, since they are plausible by itself, their functioning is derived from first principles, and they have beneficial consequences. As stated, just based on the data alone, the case to include them would not be strong enough. The result can be seen in Fig. 17.

**Fig 17.**
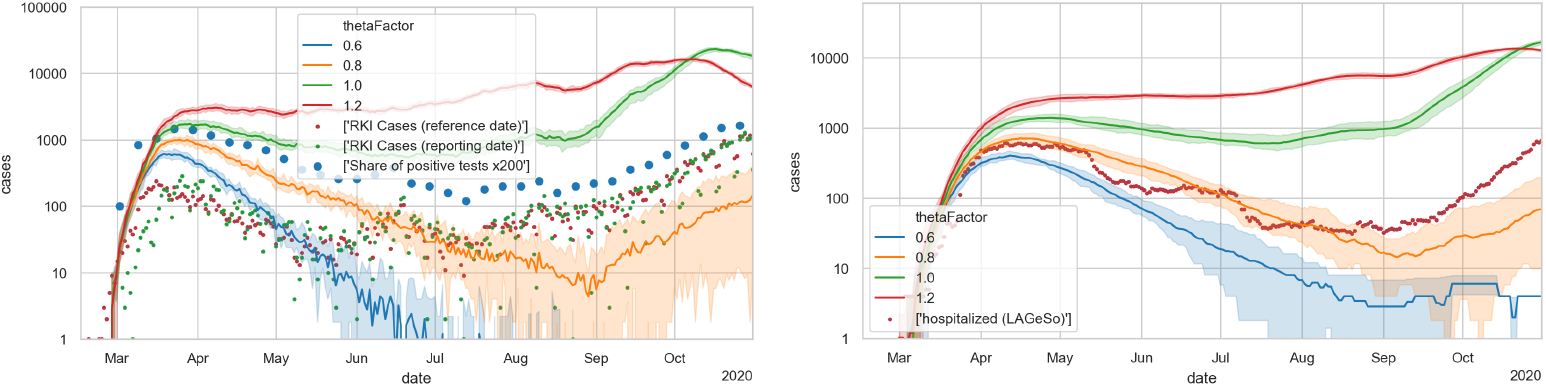
Case numbers (LEFT) and hospital numbers (RIGHT) in simulations that now also include masks and TTI (trace, test, and isolate).

### Summer disease import

After adding masks and contact tracing, the second wave is once more too late. Adding summer disease import pushes the curve up again (Fig. 18). Other than during the spring import, where we multiplied the RKI numbers by 4, we now divide them by two. The reason is that the disease import stems from the case numbers, and as can be seen in the figure, the factor between the case numbers and the re-scaled positive test fraction is much smaller in summer than in spring (also cf. Sec. Under-reporting).

**Fig 18.**
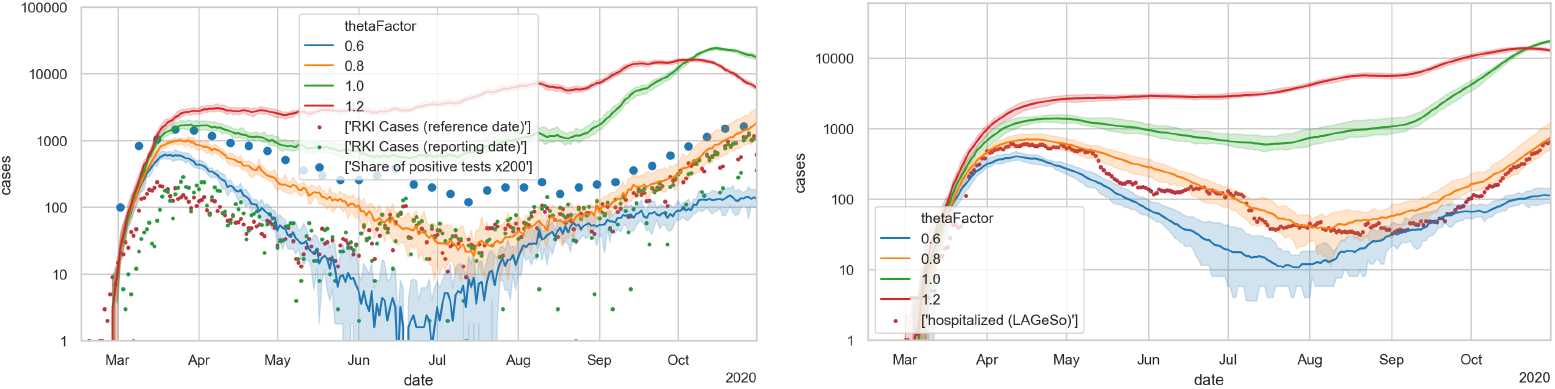
with summer import

Setting the disease import to one, as would be plausible by this argument, leads to an influence that is too large. Since there was widespread screening and an obligation to remain into quarantine-at-home for many people returning from summer travel, we argue that (1) the testing-and-quarantine regime had the consequence that many disease imports did not reinfect others and that (2) the screening also found many asymptomatic cases that would otherwise not have been included into the case numbers.

Again, the case for adding this element, and in this way, is not very strong. An alternative would have been to stay with the model of Fig. 15, with a thetaFactor between 0.6 and 0.8 (look in particular at the hospital cases). Again, we prefer adding masks, contact tracing, and summer disease import, since the models can be constructed from first principlesand as a package, they allow for a slightly larger thetaFactor, which overall seems plausible.

## Availability of data and materials

For computer code see https://github.com/matsim-org/matsim-episim. Simulations were computed with version d16656f076640124de0361fc327d3803a80aa466 of the code, started with command

~~~
java -jar matsim-episim-1.0-SNAPSHOT.jar runParallel \
 --setup org.matsim.run.batch.BerlinSensitivityRuns \
 --params org.matsim.run.batch.BerlinSensitivityRuns$Params
~~~

The input data (including the synthetic mobility traces) are made public here: https://svn.vsp.tu-berlin.de/repos/public-svn/matsim/scenarios/countries/de/episim/openDataModel/input/

The output data used for the figures can be retrieved at https://svn.vsp.tu-berlin.de/repos/public-svn/matsim/scenarios/countries/de/episim/battery/2021-02-09/.

We would be happy to put the data into a more permanent location if desired; unfortunately, our university repository currently has problems with such large data sets as ours.

## Supporting information

### S1 Text. Senozon method

Since we are interested in trajectories for large population samples, we start from the network-based approach. Because of privacy restrictions in Germany, the trajectories, even when cut into 8-hour segments, cannot be used directly. In order to be compliant with existing privacy regulations in Germany, they are therefore processed before they can be used for scientific work. The steps are [113]:

1. First, a synthetic population with attributes home location, age, gender, and employment status is generated based on available census data.
2. Separately, the signalization records, i.e. of celltower handovers, are converted into plausible movement trajectories. This step attempts to remove celltower handovers that occur because of operator load rebalancing rather than user movements, and to remove celltower handovers that occur during travel (since the method is only interested in activities).
3. The resulting trips between activities are exported as plain hourly origin-destination matrices.
4. At the same time, the movement trajectories from step 2 are annotated with demographic categories from the Customer Resource Management of the cellhphone company, and with plausible activity types, based on land-use properties of the celltower areas and time-of-day. At the same time, the locations are removed. The result, i.e. activity patterns including their times together with each pattern owner’s home zone, gender, and age category, is exported as well.
5. For each synthetic person from step 1, at least 30 patterns from step 4 are selected, based on closeness between attributes of the synthetic person and attributes of the pattern owner; one of the patterns is selected randomly.
6. The resulting activity pattern is enriched with location information for all non-home activities based on the travel times between activities and the origin-destination matrix from step 3.

Our second data set, which contains the reductions of activity participation data for each day since 2020-03-01 (cf. Figs. 12 and 2), is derived in the same way, but the computation is stopped after step 2, and only the average times spent at each activity type are aggregated per zip code and then exported.

### S2 Text. Details of the mobility model

#### Handling of large facilities

The resolution of our input data comes at the level of “facilities”. Those can be interpreted as buildings or sometimes blocks. They often contain multiple households, multiple company offices, multiple leisure facilities, multiple shops, etc. For *home* activities, we split persons living in the same facility into realistic household sizes with a maximum number of six people per household [114]. This seems important since the within-household dynamics of COVID-19, and in particular the fact that the secondary attack rate in households seems to be far below 100%, plays an important role (e.g. [115]). For all other activities, we divide the facilities by some globally set factor, called *N*^*spacesPerFacility*^. That is, if two persons spend overlapping time at the same facility, the probability that they have interacted is 1*/N*^*spacesPerFacility*^. This has important ramifications for multi-day modelling and mixing, see below.

#### Multi-day modelling

Optimally, one would have multi-day trajectories. In our case, the data that we have ends at the end of the day. Our simulations thus run the same person trajectories again and again (except for weekends, see below). This presumably *underestimates* mixing, since it is plausible to assume that there is some variation in activity patterns from day to day. At this point, one needs to make a decision whether our sub-spaces (see above) are frozen, meaning that the same sub-groups meet every day, or not. Using the same sub-groups every day arguably is plausible for office buildings, which may contain offices for several companies, and interaction may be limited to sharing an elevator. It is less plausible for public transport trains, where passengers are arranged differently every day. Possibly, a mix between the two approaches is plausible, introducing the need for even more free parameters. In our present model, we opt for the non-frozen setting, i.e. the other persons within a facility that an ego person interacts with are randomly re-drawn for every new simulated day. *N*^*spacesPerFacility*^ evidently influences the number of contacts that a person has. For our simulations, we set it such that that number of contacts is roughly consistent with real-world contact tracing. For our current input data, that leads to a setting of *N*^*spacesPerFacility*^ = 20.

#### Weekend modelling

As already alluded to above, we use separate models for Saturdays and Sundays. They come out of transport modelling in the same way as we obtain the model for a “typical weekday” (see above). These models use the same synthetic persons and facilities, and thus can be aligned with the weekday model. In consequence, each synthetic person in our models, starting on Monday, (a) repeats the same weekday five times, (b) runs her Saturday schedule, (c) runs her Sunday schedule, and then starts over.

#### 25% sample

For computational reasons, we use a 25% sample of the full population. The sample is constructed by choosing 25% of all persons in the population randomly and retaining their full trajectories. The splitting of households as described above is done *after* the sampling, meaning that we have realistic household sizes in the 25% scenario but consider only 25% of them; also, the number of contacts to determine the parameter *N*^*spacesPerFacility*^ (see above) is determined for the 25% model. We have also run the full 100% model to check that there are no major differences. The 25% model allows to finish runs within a single-digit number of hours, which was and is important for fast model turn-around driven by the the necessity for fast progress given the demand for the results by the decisionmakers. All results are reported after upscaling to 100%.

## Acknowledgements

We thank Kai Martins-Turner and Dominik Ziemke for discussions. We are grateful to BVG (Berlin public transit operator) for providing the mask compliance rates which they surveyed on a daily basis. The work on the paper was funded by the Ministry of research and education (BMBF) Germany (01KX2022A) and TU Berlin; regular reports can be found through this search: https://depositonce.tu-berlin.de/simple-search?query=modus-covid. Zuse Institute Berlin (ZIB) provided CPU time.

## Footnotes

1 https://www.statistik-berlin-brandenburg.de/BasisZeitreiheGrafik/Bas-Schulen.asp?Ptyp=300&Sageb=21001&creg=BBB&anzwer=5

2 Cf. https://depositonce.tu-berlin.de/simple-search?query=modus-covid The ANSI (1997) band-importance functions (BIFs) were interpolated.

3 Again, see https://depositonce.tu-berlin.de/simple-search?query=modus-covid.

4 A thetaFactor of 1.0 corresponds to Θ = 0.000561.

5 https://covid-sim.info/2020-11-09/tracing

